# Leveraging Pre-Vaccination Antibody Titers across Multiple Influenza H3N2 Variants to Forecast the Post-Vaccination Response

**DOI:** 10.1101/2024.08.01.24311325

**Authors:** Hannah Stacey, Michael A. Carlock, James D. Allen, Hannah B. Hanley, Shane Crotty, Ted M. Ross, Tal Einav

## Abstract

Despite decades of research on the influenza virus, we still lack a predictive understanding of how vaccination reshapes each person’s antibody response, which impedes efforts to design better vaccines. Here, we combined fifteen prior H3N2 influenza vaccine studies from 1997-2021, collectively containing 20,000 data points, and demonstrate that a person’s pre-vaccination antibody titers predicts their post-vaccination response. In addition to hemagglutination inhibition (HAI) titers against the vaccine strain, the most predictive pre-vaccination feature is the HAI against historical influenza variants, with smaller predictive power derived from age, sex, BMI, vaccine dose, the date of vaccination, or geographic location. The resulting model predicted future responses even when the vaccine composition changed or a different inactivated vaccine formulation was used. A pre-vaccination feature ‒ the time between peak HAI across recent variants ‒ distinguished large versus small post-vaccination responses with 73% accuracy. As a further test, four vaccine studies were conducted in 2022-2023 spanning two geographic locations and three influenza vaccine types. These datasets formed a blinded prediction challenge, where the computational team only received the pre-vaccination data yet predicted the post-vaccination responses with 2.2-fold error, comparable to the 2-fold intrinsic error of the experimental assay. This approach paves the way to better utilize current influenza vaccines, especially for individuals who exhibit the weakest responses.

## Introduction

Although the number of potential influenza vaccine designs has rapidly grown over the past years, we lack a clear picture of which individuals will poorly respond to a vaccine or what modifications would improve outcomes. Currently, vaccine design is “virus-centric,” where the primary goal is to ensure the vaccine strain (or variant) matches the dominant circulating strain in the coming season.^1–3^ While these efforts are complemented by ferret studies (infecting ferrets with historical strains to assess how they inhibit current strains), these ferret data do not capture the heterogeneity nor the complex immune history in humans, leading to a disconnect between vaccine strain selection and the subsequent response it elicits.^3–5^ Indeed, influenza vaccine effectiveness remains around 20-50% even in seasons when the vaccine strain matches the dominant circulating strain, with some individuals showing no measurable response to the vaccine.^6–9^ This underscores the need for a combined virus-and-people-centric approach based upon both a strain’s prevalence and the immunity it elicits in people.

Vaccination induces a complex cascade of immune interactions spanning innate and adaptive immunity.^5^ In this work, we focus on the antibody response that plays a central role in mediating protection against influenza, specifically focusing on the hemagglutination inhibition (HAI) assay that correlates with protection.^10–12^ HAI quantifies how antibodies within sera block the virus from binding sialic acids on red blood cells, with higher HAI titers associated with greater protection.^13^

Although numerous studies use HAI to assess vaccine responses, we lack frameworks that can predict each person’s vaccine response *a priori* and identify why the same vaccine elicits a strong HAI response in some but little-to-no response in others (**Fig 1A**). Prior exposures may boost subsequent vaccine responses^14,15^ or prevent development of *de novo* B cell responses.^16,17^ Antigenic seniority, imprinting, vaccine blunting, and antibody ceiling effects have been observed, often at the population level, yet it is unclear how they weave together to shape each person’s post-vaccination response.^6,18–21^

**Figure 1.**
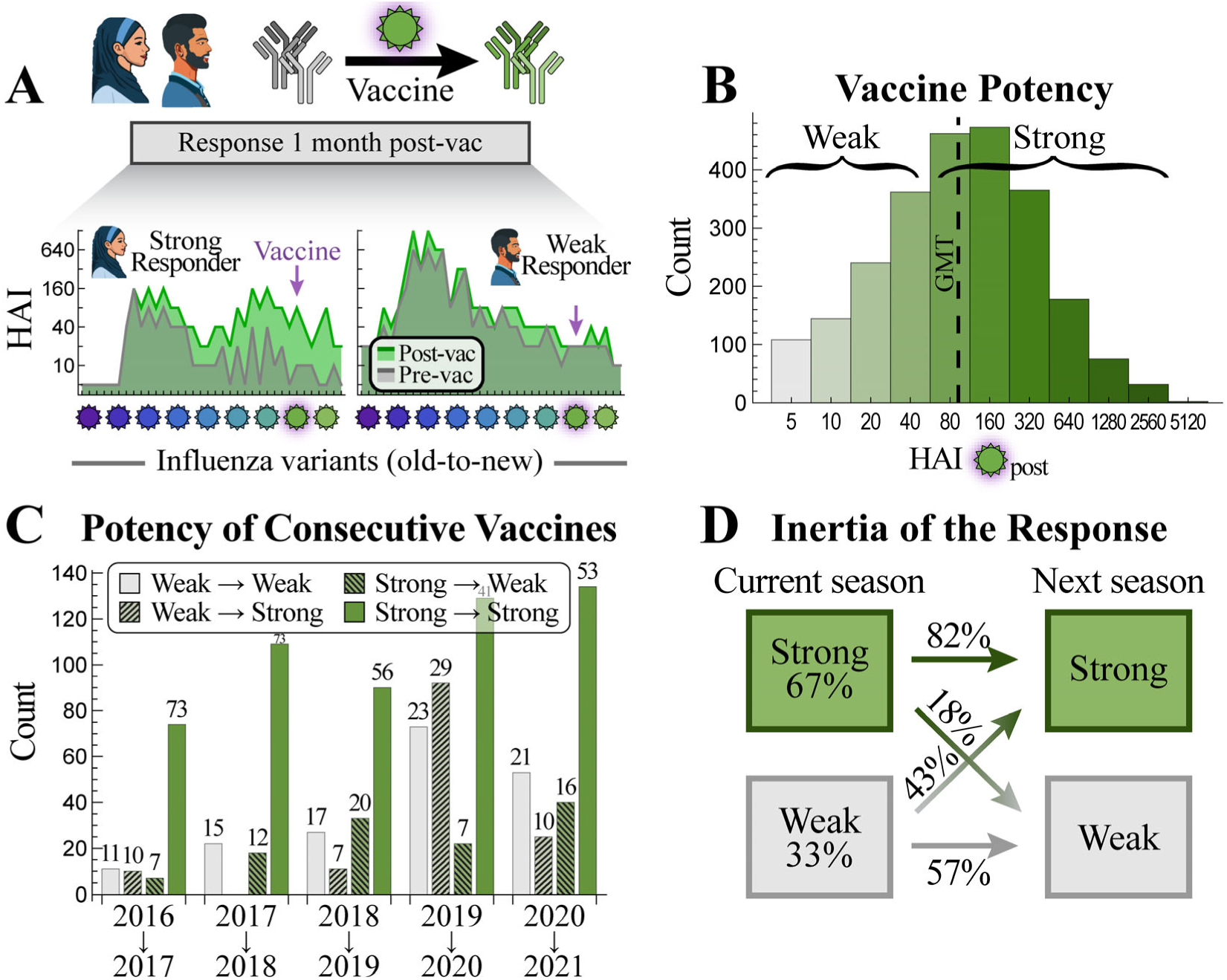
Characteristics of the vaccine response across multiple studies. (A) Antibody inhibition against multiple variants is heterogeneous across the population. Two representative examples of a strong and weak vaccine response (IDs 69 and 117 from the 2016 Fox_HCW_ dataset, **Table 1**) showing pre-vac [gray] and 1-month post-vac HAIs [green]. The vaccine strain is denoted by a pink halo and variants are ordered from oldest-to-newest [purple-to-green viruses; past variants to the left of vaccine strain, future variants to its right]. (B) Distribution of vaccine strain HAI titers 1-month post-vac across the 15 prior studies from **Table 1**. The HAI geometric mean titer (GMT=95) is shown as a dashed line. (C) Tendency of vaccine responses to be strong (HAI≥80) or weak (HAI≤40) over two consecutive seasons (*x*-axis, all from the UGA studies). The percent in each class is shown above the bars. (D) Summary of strong or weak responses across all years (boxes) and the tendency to maintain these profiles over consecutive seasons (arrows).

**Table 1.** List of large-scale influenza vaccine studies. Fifteen existing vaccine studies (white background) were used to train and test the model forward in time. Four new vaccine studies (gray) were exclusively used to test the model. The total number of measurements in each study equals (# of sera)×(# of viruses)×(2 time points [pre– and post-vac]).

| Year + Name of Study* | # of Measurements | # of Sera | # of Viruses | Vaccine Type |
| --- | --- | --- | --- | --- |
| 1997 Fonville<sup>4</sup> | 7420 | 106 | 70 | Inactivated (Afluria trivalent) |
| 1998 Fonville<sup>4</sup> | 8960 | 128 | 70 | Inactivated (Afluria trivalent) |
| 2009 Fonville<sup>4</sup> | 1600 | 80 | 20 | Inactivated (Afluria trivalent) |
| 2010 Fonville<sup>4</sup> | 1600 | 80 | 20 | Inactivated (Afluria trivalent) |
| 2014 Hinojosa<sub>V</sub><br>(Vaccinated or infected)<sup>37</sup> | 656 | 41 | 16 | Inactivated (Fluzone trivalent) or<br>Live attenuated (FluMist quadrivalent) |
| 2015 Hinojosa<sub>V</sub><br>(Vaccinated or infected)<sup>37</sup> | 656 | 41 | 16 | Inactivated (Fluzone trivalent) |
| 2015 Hinojosa<sub>U</sub><br>(Unvaccinated and uninfected)<sup>37</sup> | 496 | 31 | 16 | Inactivated (Fluzone trivalent) |
| 2016 Fox<sub>Nam</sub><br>(Ha Nam)<sup>38</sup> | 4100 | 100 | 41 | Inactivated (Vaxigrip trivalent) |
| 2016 Fox<sub>HCW</sub><br>(Health Care Workers)<sup>38</sup> | 1764 | 49 | 36 | Inactivated (Fluarix quadrivalent) |
| 2016 UGA<sup>39</sup> | 2516 | 148 | 17 | Inactivated (Fluzone trivalent) |
| 2017 UGA<sup>39</sup> | 4878 | 271 | 18 | Inactivated (Fluzone trivalent) |
| 2018 UGA<sup>39</sup> | 2500 | 250 | 10 | Inactivated (Fluzone trivalent) |
| 2019 UGA<sup>39</sup> | 3688 | 461 | 8 | Inactivated (Fluzone trivalent) |
| 2020 UGA<sup>39</sup> | 2373 | 339 | 7 | Inactivated (Fluzone quadrivalent) |
| 2021 UGA<sup>39</sup> | 2359 | 337 | 7 | Inactivated (Fluzone quadrivalent) |
| 2022 UGA | 175 | 25 | 7 | Inactivated (Fluzone quadrivalent) |
| 2023 UGA | 150 | 25 | 6 | Inactivated (Fluzone quadrivalent) |
| 2023 Crotty<sub>Afluria</sub> | 168 | 24 | 7 | Inactivated (Afluria quadrivalent) |
| 2023 Crotty<sub>FluMist</sub> | 175 | 25 | 7 | Live attenuated (FluMist quadrivalent) |

\* Each year represents the date when a vaccine study was conducted, not when the corresponding manuscript was published.

In addition, various host features (*e.g.*, age,^18,22,23^ genomics,^24,25^ pre-vac HAI^26,27^) have been shown to affect the antibody response. These effects are often assessed one study at a time, and it is unclear whether they are universal or study-specific. Here, we demonstrate how combining multiple studies readily quantifies how any combination of parameters constrains the post-vac response (without requiring any modeling). The most predictive variables provide a roadmap for the key features to measure in future studies, as well as providing the ingredients for modeling the vaccine response.

Using those maximally predictive variables, we develop an algorithm that takes a person’s HAI titers pre-vaccination (pre-vac) to predict their peak HAI response 3-4 weeks post-vaccination (henceforth referred to as “1-month post-vac”), focusing exclusively on the H3N2 subtype. Our key findings include: (1) By combining data from prior vaccine studies, heterogeneous vaccine responses across ≥10 influenza seasons (from 2009 to 2021) can be predicted with accuracy comparable to experimental noise. (2) Prediction accuracy holds across four new vaccine studies we conduct (in 2022 and 2023) spanning three vaccine types and two geographic locations. For this challenge, the computational team (T.E.) was blinded and only given the pre-vac data to stringently test the model’s predictive power. (3) The most informative variables for the post-vac response are an individual’s pre-vac HAI against the vaccine strain, their pre-vac HAI against other influenza variants, whether they receive an inactivated versus live attenuated vaccine, and their prior influenza exposure history. (4) While individual studies may find different relationships between variants, combining all studies from the past decade leads to universal relations that accurately predict post-vac titers. (5) The magnitude of the fold-change post-vac is strongly associated with the # of years (Δ_Peak_) between the two most recent peaks in the HAI landscape; 2≤Δ_Peak_≤3 yields a large fold-change while 4≤Δ_Peak_≤6 leads to a smaller fold-change in 73% of cases.

The resulting algorithm is built upon a machine learning approach that finds relations between pre– and post-vac HAI titers from large-scale influenza vaccine studies going back to 1997. Instead of splitting each dataset into training/testing sets, we restrict ourselves to the harder prediction challenge of training on some datasets and testing on *entirely different datasets*. Predictions are done forward in time; for example, to predict a person’s vaccine response in 2020, training was done on data from 2019 and before. This model only uses pre-vaccination measurements as input, and hence it can take any individual’s pre-vac HAI and predict their post-vac HAI, without needing to “peek” at the initial vaccine response.^4,28–32^

## Results

### The Inertia of Vaccine Responses elicits Consistently Strong/Weak Responses across Seasons, especially when the Vaccine Strain is Unchanged

We performed an extensive literature search for influenza vaccine studies from the past two decades. Due to the substantial effort involved, such studies often restrict their analyses to the sizable datasets they produce. Here, we chose the opposite tact and characterized responses across cohorts. Doing so builds towards a fundamentally new question, namely, whether all post-vaccination HAI data in each study can be entirely predicted by prior studies, even as the virus evolves and the vaccine strains change.

The literature search found 15 influenza vaccine studies measuring pre– and post-vac serum HAI in ≥25 people against ≥6 H3N2 variants (**Table 1, Fig S1**). On average, cohorts included 160 (from 31-461) sera that showcase the heterogeneity of responses, measured against 30 H3N2 variants (from 7-70). Collectively, these ∼2,500 sera measure vaccine responses from much of the past decade (2014-2021) as well as four prior years (1997, 1998, 2009, 2010), providing ample opportunities to characterize vaccine response within and across influenza seasons.

The HAI of the vaccine strain 1-month post-vac followed an approximate log-normal distribution with a geometric mean titer of GMT=95 (**Fig 1B**). Classic studies found that an HAI of 40 or 80 correlated with 50% protection against H3N2 infections.^33–36^ Adopting the conservative stance where HAI≥80 is deemed a strong response, 67% of sera had strong post-vac responses on average.

Across all studies, individuals vaccinated in two consecutive seasons showed a tendency towards maintaining their same strong or weak response, although there was a global bias towards strong responses (**Fig 1C,D**). More precisely, someone exhibiting a strong response was 82%/18%≈4x as likely to exhibit a strong response the next season, while someone with a weak response was only 57%/43%≈1.3x as likely to maintain their weak response (**Fig 1D**).

On average, 73%(=0.82ꞏ67%+0.57ꞏ33%) of individuals maintained their strong→strong or weak→weak responses across all years examined (**Fig 1D**), yet ≥83% did so during seasons when the H3N2 vaccine strain was unchanged or minimally changed (**Table S1**, Spearman’s rank test *p*=0.04 using the HA epitopes [ΔAA_Epitope_], *p*=0.02 using full HA [ΔAA_Total_]). Taken together, these results demonstrate that features of the vaccine strain and the antibody response from (at least) one prior season can help inform future responses.

### A Model-Free Approach to Quantifying Predictive Power of Host Traits

Multiple factors have been shown to affect the antibody response, including age or birth year,^18,21,40–42^ genetics,^43,44^ and a person’s vaccination or infection history.^38,45–47^ Yet the amount each factor, or a combination of factors, affects the response is harder to quantify. Powered by the 15 studies, we introduced a simple, model-free method to assess how different combinations of features constrain the post-vac HAI against the vaccine strain.

As an example, we first assessed two commonly measured features: a person’s age and their pre-vac HAI against the vaccine strain. To quantify how these two features constrain the post-vac response, every pair of individuals that have a similar age and pre-vac HAI (age ≤5 years apart, HAI exactly matches) were considered, and the ratio of their vaccine strain’s HAI 1-month post-vac was computed (which should be ≈1 if they maintain a similar response).

Given the ultimate goal of predicting vaccine responses across seasons, only pairs of subjects vaccinated in *different* years were considered to test whether features fully specify the post-vac HAI over time. For the specific case of age and pre-vac HAI, *N*≈110,000 pairs matched on both traits across all studies, and their post-vac HAI differs by 4.0x root-mean-squared error (RMSE, “x” denotes fold-change; 95% CI: 4.0-4.05x) (**Fig 2A,B** green box). By assessing each feature separately, most of this predictive power came from the pre-vac HAI (RMSE=4.5x, 95% CI: 4.47-4.5x, **Fig 2B** red). Age alone poorly constrained the post-vac response across all age groups (RMSE=5.9x, 95% CI: 5.88-5.91x, **Fig 2B** gold), although exclusively considering ages≥65 led to more homogeneous responses and slightly better prediction accuracy (**Fig S2**).

**Figure 2.**
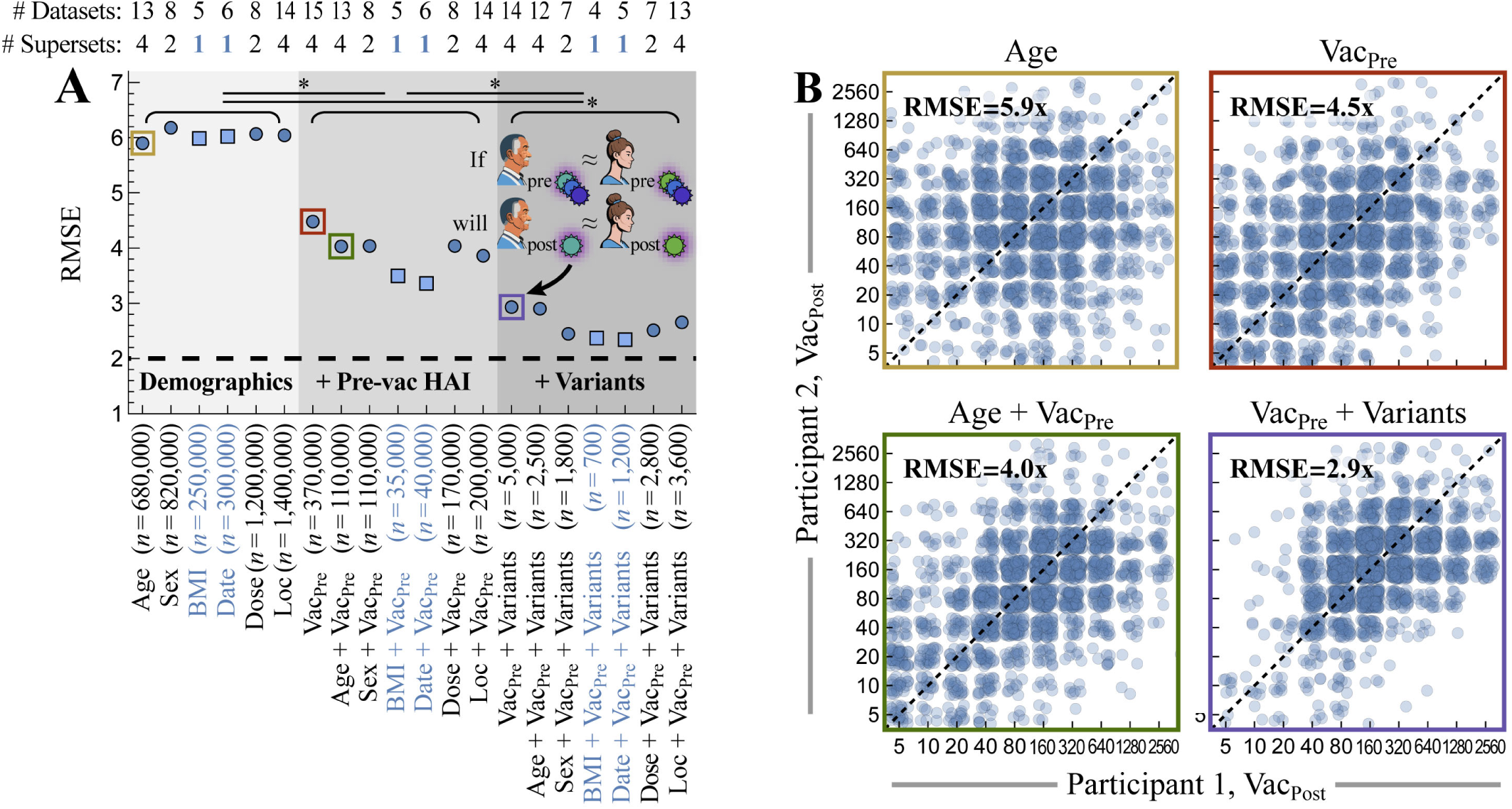
A model-free way to identify the key parameters that predict vaccine responses across studies. (A) For all pairs of subjects vaccinated in different seasons that match across a feature set [*x*-axis], their post-vac HAI was compared against their respective vaccine strains. Features include body mass index (BMI), the date of vaccination (Date), standard vs high dosage (Dose), geographic location (Loc), pre-vac HAI against the vaccine strain (Vac_Pre_), and pre-vac HAI against other variants (**Methods**). Root-mean-squared error (RMSE, *y*-axis) is shown for each feature set, assessed over *n* pairs of subjects. 95% CI are smaller than the plot markers. Blue circles indicate that pairs of matching individuals can be found across multiple supersets; teal squares indicate that matched pairs may have artificially small RMSE because they came from a single superset of studies (UGA, Fox, Hinojosa, or Fonville; all cases of 1 superset came from UGA studies). The dashed line represents the intrinsic 2-fold error of the HAI assay.^4^ \**p*<0.001 between any feature sets with different gray shading (*e.g.,* Age vs Vac_Pre_ or Date vs Vac_Pre_+Variants) using a one-sided permutation test. (B) Vaccine strain’s post-vac HAI (Vac_Post_) across all pairs matching on four potential feature sets. For clarity, at most 2000 (randomly selected) points are shown, although statistics were computed on all data.

We similarly assessed other combinations of features measured in the 15 studies, which included a participant’s age, sex, BMI, their date of vaccination, dosage, geographic location, pre-vac HAI against the vaccine strain, and pre-vac HAI against other variants (**Methods**). Among these features, the largest decrease in RMSE occurred when matching the pre-vac HAI against the vaccine strain, resulting in a significant decrease to RMSE regardless of whether other features are added (*p*<0.001, one-sided permutation test, **Fig 2A**). After matching pre-vac HAI against the vaccine strain (**Fig 2A** red square), further matching pre-vac HAI across all variants led to a further significant decrease to RMSE (*p*<0.001, one-sided permutation test, **Fig 2A** purple square). Other features, ordered from most-to-least predictive power, were the date of vaccination, BMI, geographic location, age, sex, and vaccine dose, each providing smaller but still significant decreases (*p*<0.001, one-sided permutation test, **Fig 2A)**.

Note that BMI and the date of vaccination were only measured in the UGA studies. Thus, any set including these features (shown by teal squares in **Fig 2A**) may have artificially lower error, since these cohorts necessarily used the same study design, administered the same vaccine, and were carried out in the same geographic location. Indeed, combining BMI or the date of vaccination with the vaccine’s HAI slightly decreased RMSE, although such results should be reassessed against future vaccine studies from other groups. All other features were measured in more than one superset (defined as UGA, Fox, Hinojosa, or Fonville, top of **Fig 2A**), leading to more robust estimates with subject pairs that span across study designs.

### Forecasting the Vaccine Response

We next determine how well pre-vac HAIs predict the post-vac response, while taking into account the heterogeneity of responses, the different variants measured in each study, and differences in study design that may affect the response. Predictions were built upon a series of random forests, with one forest created to predict each variant’s post-vac HAI based on every prior study with ≥4 overlapping variants (with these variant’s pre-vac HAI used as model input). In total, ∼10^3^ separate models were collectively built to forecast the vaccine response (**Methods**).

Rather than splitting each dataset into a training and testing set ‒ which for future application would require some individuals to get vaccinated early each season to predict others’ responses ‒ we instead strive for the tougher challenge of training exclusively on vaccine studies from prior years. Since many influenza studies (beyond the ones analyzed in this work) only measure the vaccine strain, we sought to beat the 4.5x error found when only matching the vaccine strain’s pre-vaccination HAI (Vac_Pre_, **Fig 2A** red square), ideally aiming for the 2x noise limit of the HAI assay (see **Methods** for the quantification of assay noise). We also compare model performance against a null model (assuming no HAI response) and a linear model (assuming a linear response for each variant).

### Equating Analogous Variants across Studies

Since predictions were based on the HAI of overlapping variants between studies, we increased this overlap by equating variants whose HA sequences differed by ΔAA_Epitope_<5 amino acids in H3N2 epitopes A-E. This threshold balanced the twin goals of equating more strains while ensuring that equated strains have similar HAI profiles (**Fig S3A**). For example, HAI from H3N2 A/Hong Kong/1/1968 ≈ A/Bilthoven/16190/1968 ≈ A/Aichi/2/1968 were combined for model training and testing (**Fig 3A**; full list in **Fig S3**). In total, this decreased the total number of unique variants across all studies from 102→80.

**Figure 3.**
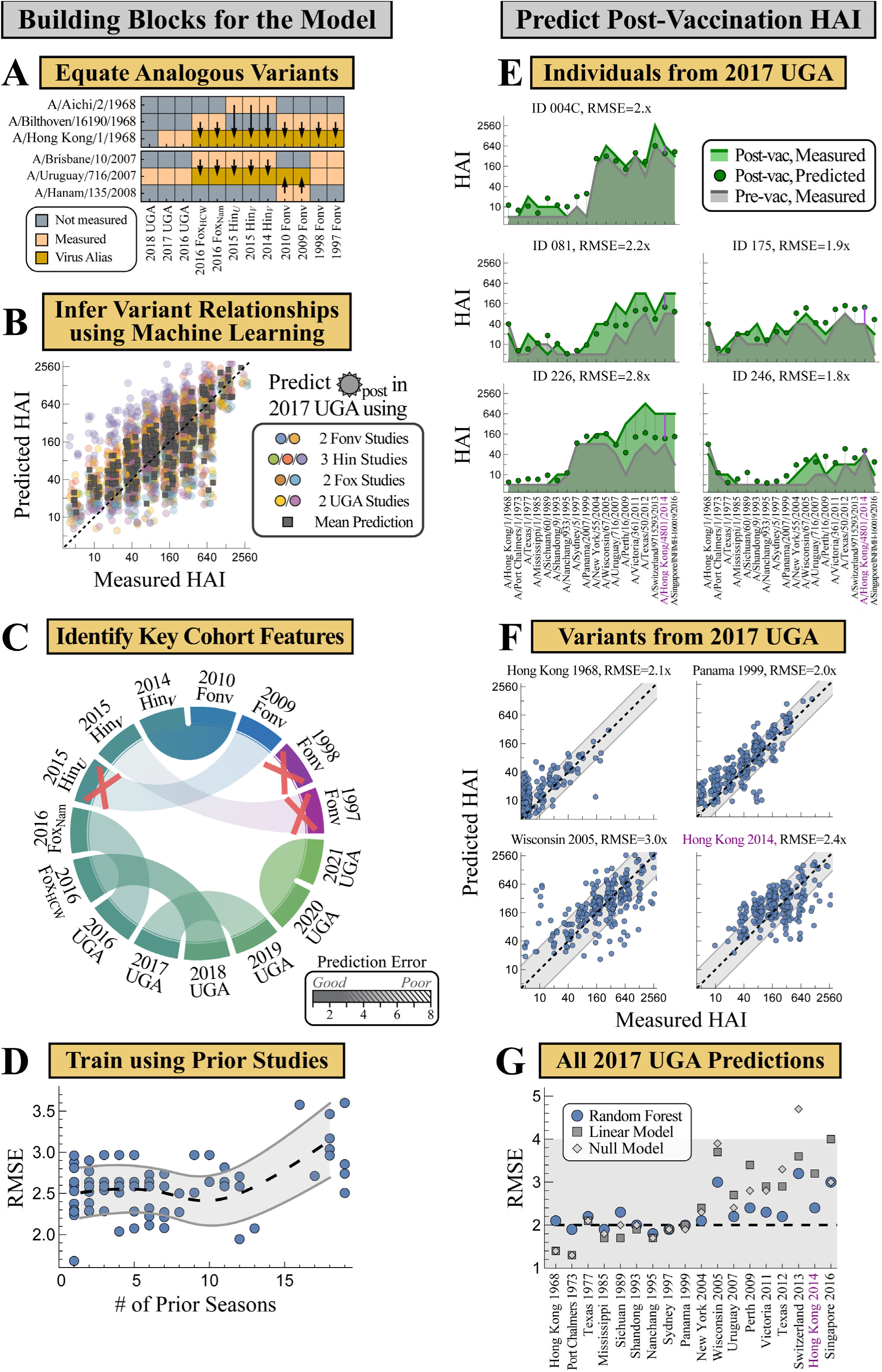
Using pre-vaccination HAI against multiple variants to predict HAI 1-month post-vaccination. (A) Analogous variants with similar HA sequences are equated (full list in **Fig S3**). Pre-vac HAI from these overlapping variants served as model input to predict post-vac HAI. (B) Example predictions for H3N2 A/Uruguay/716/2007 in 2017 UGA using 2009-2010 Fonville, all Hinojosa and Fox studies, 2016 UGA, and 2018 UGA (circles), together with the geometric mean across all 9 studies (squares). Each point represents one subject. (C) Representative chords showing prediction RMSE across pairs of studies (full chords in **Fig S5**). The three crossed-out studies have upper quartile RMSE>4x. (D) Prediction error from training on all vaccine studies from the prior *n* seasons [*x*-axis]. Mean [black dashed] and standard deviation [gray lines] of these RMSEs are interpolated across *n*. (E) Example predictions of five individuals in 2017 UGA [subject IDs and RMSE across variants shown above]. Pre-vac HAIs [gray line] predicted the post-vac response [green points], and results were compared to post-vac data [green line]. Prediction error is emphasized by the vertical lines [purple bars and text highlight the vaccine strain H3N2 A/Hong Kong/4801/2014, also shown in the bottom-right of Panel F and in Panel G]. Variants are sorted from oldest to newest (left-to-right). (F) Predicted versus measured post-vac HAI of four variants across all 271 subjects in 2017 UGA. The diagonal line *y*=*x* represents perfect predictions, while the gray bands denote the RMSE of the predictions. (G) RMSE for all variants in 2017 UGA comparing the random forest approach (blue points) with a linear model (gray squares) or null model (gray diamonds). The gray region denotes predictions with the desired RMSE≤4x.

All subsequent analysis was carried out after equating these analogous strains. In all cases where virus analogues were measured in the same study, their HAI titers were nearly identical, as expected (**Fig S3**). However, prior work has shown that some single substitutions lead to dramatically different HAI reactivity,^48^ and hence the ultimate test for these equivalences will be the accuracy of the final forecasts.

### Inferring Global Relationships between the Pre– and Post-vaccination HAI of Influenza Variants

To predict the post-vac HAI for virus-of-interest *V*_0_ in study *Y* using data from study *X* (also measuring *V*_0_), a prior random forest algorithm^31^ was modified to account for the pre– and post-vac time points. Briefly, five randomly chosen viruses measured in both studies (*V*_1_-*V*_5_, at least one must be *V*_0_) were used to train decision trees using *pre*-vac HAI from *V*_1_-*V*_5_ as input to predict the *post*-vac HAI of *V*_0_ as output (**Methods**). 50 trees were created, each internally trained on 30% of sera from study *X* and tested on the remaining 70%. The top 5 trees comprised the final random forest model, with the average of their HAI outputs serving as *V*_0_‘s predicted post-vac titers in study *Y*.

In essence, this algorithm searched for the most predictive overlapping variants for *V*_0_. For example, the HAI of H3N2 A/Uruguay/716/2007 in the 2017 UGA cohort could be predicted by nine other vaccine studies that measured this same variant, with each study estimating the post-vac HAI for all 271 individuals in the 2017 UGA study (**Fig 3B**). Taking the geometric mean of all nine values as the final prediction led to an RMSE=2.3x (95% CI: 2.1-2.4x), comparable to the ≈2x error of the HAI assay. While pre-vac HAI for Uruguay 2007 was among the top two features for most trees, other historical or future variants had comparable importance (**Fig S4**).

### Using Poor Predictions to Identify the Factors that Affect Post-Vaccination HAI

This compilation of random forests predicted the post-vac HAI of all overlapping variants between any pair of studies. Although we ultimately desired accurate predictions, poor estimates revealed which features fundamentally affect the antibody response in a complementary manner to **Fig 2**.

For each pair of vaccine studies, post-vac HAIs were predicted for all overlapping variants, with the least accurate predictions depicted with more transparency (a few chords shown in **Fig 3C**, all chords in **Fig S5**). While predictions were generally accurate, three datasets exhibited systematically poor predictions (with upper quartile RMSE>4x, **Fig S5**).

The two earliest studies (1997-1998 Fonville) poorly predicted all subsequent vaccine studies (from 2009 or later). Unsurprisingly, this suggests that as antibody responses change over time, prior responses poorly predict outcomes too far into the future. This effect will be examined systematically in the following section.

The only other study with uniformly poor predictions was 2015 Hinojosa*_U_*, a study comprising children that self-reported as being uninfected and unvaccinated to influenza during the past five years. Notably, the 2014 and 2015 Hinojosa*_V_* vaccine studies (children that were either infected or vaccinated in the past five years) had uniformly accurate predictions (interquartile RMSE between 2-3x, **Fig S5**), demonstrating that children can predict adult responses and vice versa. Instead, these results suggested that the unique exposure history of the children in 2015 Hinojosa*_U_*led to their fundamentally different antibody responses. Since this was the only study with such exposure histories, it was dropped in the subsequent analysis. All remaining studies had upper quartile RMSE<4x, so we next turn to the final step of determining how many prior seasons should be used to forecast future responses.

### This Season’s Vaccine Responses are Informed by Responses from the Past 10 Seasons

The above framework gave pairwise predictions from study *X*→*Y*. While predictions from a combination of studies (*X*_1_, *X*_2_…)→*Y* will likely be more robust, the number of possible training sets grows combinatorially; for example, the 2020 UGA study can be predicted using any combination of the 13 prior datasets (2^13^≈8,000 choices).

For each study, the average post-vaccination response was computed from all vaccine studies conducted between 1 and *n* seasons beforehand (**Fig 3D**). Prediction accuracy was maintained while using *n*≤10 prior seasons, so the final algorithm combined the predictions of all vaccine studies from the past decade to forecast future antibody responses.

### Post-vaccination HAI is Predicted with 2.4-Fold Error across 15 Vaccine Studies

The above framework uses a person’s pre-vac HAI to predict their response 1-month post-vac. As an example, forecasts of the 2017 UGA vaccine study were assessed against the measured responses (results for all studies in **Fig S6**). Pre– and post-vac titers are shown for 5 (of 271) individuals, demonstrating the heterogeneity in both baseline HAI and the vaccine response (**Fig 3E**). The RMSE across all 271 individuals is 2.3x (95% CI: 2.2-2.3x), comparable to the intrinsic noise of the HAI assay, and hence these predictions are as accurate as should be possible (**Fig 3E**, IDs 004C, 175, 246, 081).

Notably, some individuals showed large consistent deviations (**Fig 3E**, ID 226). To quantify this trend, predictions were extended across all studies to compute the deviation of the 5 worst-predicted variants for each serum. Overall, 9% of serum responses were substantially underpredicted (predictions ≥4x below measurements for 5 worst-predicted variants on average) and 3% were overpredicted (predictions ≥4x above measurements), while the remaining 88% of sera showed consistently small deviations across all variants (examples in **Fig S6**).

In addition to assessing each person’s response, a “transposed” analysis quantified each variant’s predictions. Returning to 2017 UGA, both the predicted and measured post-vac HAIs against the vaccine strain H3N2 A/Hong Kong/4801/2014 ranged from 40 to 640, resulting in a 2.4x prediction error (**Fig 3F**, purple bars in **Fig 3E** denote five such points). Post-vac HAIs were smaller for older variants, yet the distribution of titers showed a strong tendency to lie on the diagonal line that denotes accurate predictions (**Fig 3F**, remaining variants in **Fig S6**).

Considering all variants in 2017 UGA, the RMSE for 15/18=83% were comparable to the ≈2x noise limit of the HAI assay (**Fig 3G**). The final 3 variants had a larger RMSE≈3x, still within the target range of ≤4x. Extending this analysis to all vaccine studies, 90% of variants were predicted with RMSE≤3x while the remaining 10% were predicted with 3x<RMSE≤4x (**Fig S6A**). Collectively, this represents 20,000 measurements with an average prediction error of 2.4x.

The relatively small vaccine responses across all studies suggested that a null model (HAI_post_=HAI_pre_) or a linear model (only using each variant’s HAI_pre_ to predict its HAI_post_) may predict these data equally well. Testing both models led to a marginally larger average prediction error of 3.0x across all 20,000 measurements, especially for more recent variants (**Fig 3G**, **Fig S7**). Notably, both models were far more prone to large outliers. Whereas *all* variants were predicted with ≤4x error with the random forest approach, 16/133=12% of variants had error>4x (from 4.4-9.8x, *p*=0.001 via a one-sided permutation test) with the null model, whereas 14/133=11% of variants had error>4x (from 4.2-52.9x, *p*=0.006 via a one-sided permutation test) with the linear model (**Fig S7, Fig S8**). Hence, the random forest approach led to slightly more accurate but far more robust predictions.

### A Prediction Challenge across Future Seasons

The above analysis was carried out on all existing datasets, with predictions done forward in time (*e.g.*, the 2020 vaccine study was predicted using the ∼40,000 measurements from 2019 and earlier, **Table 1**). To further demonstrate that prior vaccine studies can predict future responses, even in light of continual virus evolution, H3N2 responses 1-month post-vac were predicted in four new vaccine studies that we conducted (one in 2022, three from the latest 2023 season, **Fig 4A**). For this prediction challenge, the computational team (T.E.) was blinded by only receiving the pre-vac (day 0) HAIs which they used to predict the post-vac responses.

**Figure 4.**
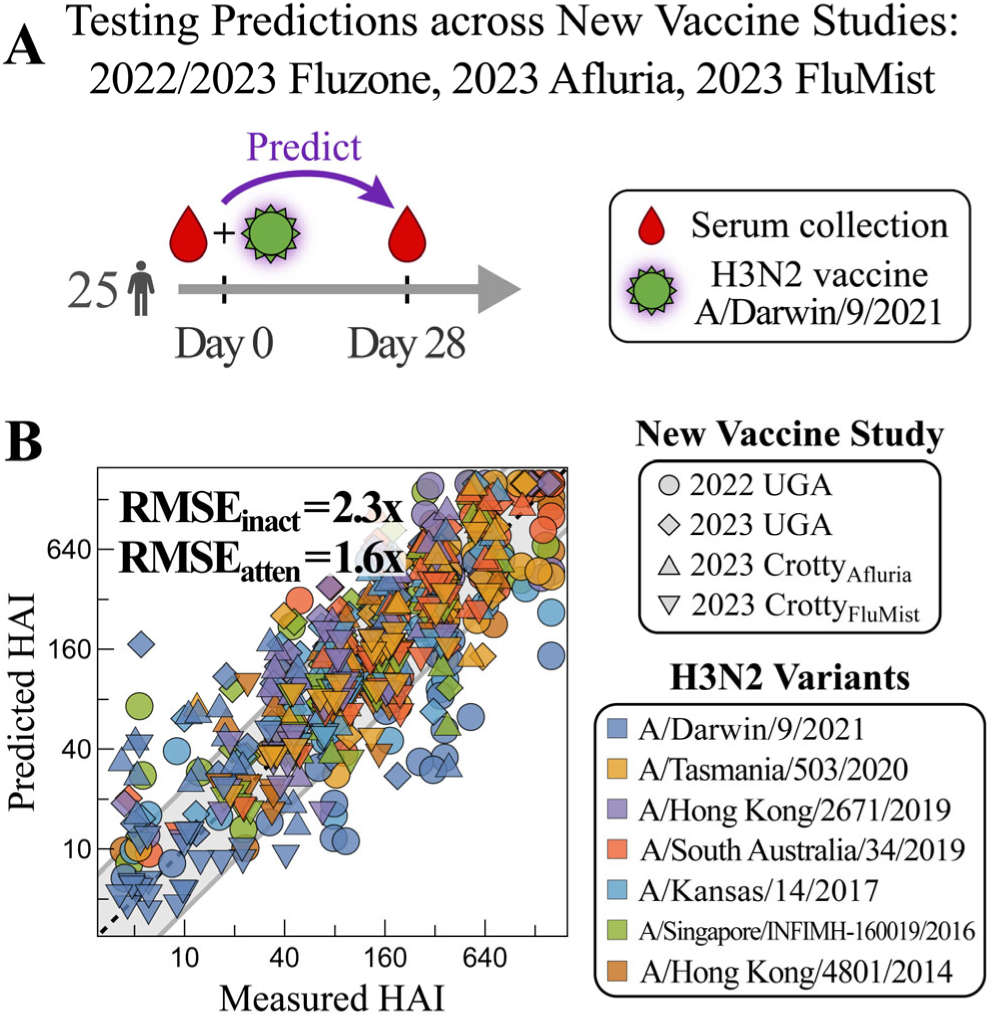
Predicting Post-vac HAI responses from new vaccine studies. (A) Four new vaccine studies were carried out administering either the 2022 or 2023 quadrivalent influenza vaccine. Each study enrolled 25 participants, and post-vac sera were collected after 28 days and measured against a panel of H3N2 variants. (B) The predicted versus measured post-vac HAIs for all variants in all four studies (individual variant predictions in **Fig S9**). Error is shown for the three inactivated vaccine studies (RMSE_inact_ for 2022-2023 UGA and 2023 Crotty_Afluria_) and the live attenuated vaccine study (RMSE_atten_ for 2023 Crotty_FluMist_). The diagonal line *y*=*x* represents perfect predictions, while the gray bands denote RMSE=2.3x for the inactivated predictions.

Two of these vaccine studies (2022 UGA and 2023 UGA) followed the same format as the prior UGA cohorts,^39^ administering the inactivated Fluzone vaccine to participants in Athens GA (demographics in **Fig S1**). The other two studies administered either Afluria or FluMist in San Diego CA (2023 Crotty_Afluria_ and Crotty_FluMist_). All four studies enrolled 25 participants and administered the seasonal vaccine containing the H3N2 vaccine strain A/Darwin/9/2021, a new vaccine introduced in 2022 and also used in 2023. Each study measured HAI titers against this vaccine strain as well as six historical variants (A/Hong Kong/4801/2014, A/Singapore/INFIMH-160019/2016, A/Kansas/14/2017, A/South Australia/34/2019, A/Hong Kong/2671/2019, and A/Tasmania/503/2020) that were chosen to overlap with the 2020 and 2021 UGA studies.

Unlike inactivated vaccines, the live attenuated vaccine FluMist has no known correlates of protection. The one other example in **Table 1** using FluMist found no increase in post-vac serum HAI.^37^ More generally, some studies have reported that live attenuated vaccines elicit little-to-no HAI responses,^49–52^ others have found HAI increases in children <18 years old,^53–56^ and a randomized double-blind clinical study found a very small HAI rise of 1.05-fold against H3N2 in adults.^57^ Since the Crotty_FluMist_ cohort comprised individuals >18 years old, predictions were made using the null model that assumed each variant’s post-vac HAI equals its pre-vac titer. In contrast, the inactivated vaccines Fluzone and Afluria were predicted using the random forest algorithm described above.

Collectively, random forests predicted the three inactivated vaccine studies [2022-2023 UGA, 2023 Crotty_Afluria_] with an average RMSE=2.3x (95% CI: 2.22-2.48x) (**Fig 4B**), outperforming both the null (RMSE=2.6x, 95% CI: 2.38-2.80x) and linear models (RMSE=2.6x, 95% CI: 2.46-2.79x). The largest prediction error occurred for the vaccine strain Darwin 2021 (RMSE=2.7‒ 4.4x), while the remaining variants had ≲2x error (**Fig S9**).

Live attenuated predictions for 2023 Crotty_FluMist_ were better for the null model (RMSE=1.6x, 95% CI: 1.48-1.68x) than the linear model (RMSE=2.3x, 95% CI: 2.17-2.49x, **Fig S9**). The null model was consistently within the noise threshold of the HAI assay (<2x), confirming that HAI titers do not noticeably increase in adults receiving this live attenuated vaccine.

As before, each vaccine study was predicted forward in time (*e.g.*, models of the 2023 UGA and 2023 Crotty_Afluria_ vaccines were trained on vaccine studies from 2022 and earlier). However, since the 2023 UGA study was carried out in September while the 2023 Crotty_Afluria_ study was done in December, the former study could augment the latter’s predictions. Rather than training on all vaccine studies from the last 10 years, models were trained using every subset of these studies, and the subset yielding the best predictions on 2023 UGA was used to predict the 2023 Crotty_Afluria_ responses. These specialized predictions led to a small but not significant improvement in the Darwin 2021 vaccine strain predictions (RMSE=3.5→3.0x, *p*=0.2 from one-sided permutation test) and minimally improved the other variants (**Fig S9E**).

### Using the Model to Recoup Known Phenomena and Discover Predictive Features of Vaccine Responses

The model provides an opportunity to search for general relationships between an individual’s pre– and post-vaccination HAI. Few such rules are known, with one major exception being the antibody ceiling effect where the strongest pre-vac responses hit a “ceiling” and exhibit smaller fold-change post-vac. Other results have been noted in individual studies; for example, the 2016 Fox_Nam_ study found that individuals recently infected by H3N2 have stronger post-vac HAI than those with no recent exposure.^38^

Both effects are captured to an extent by the model. Recently infected subjects in 2016 Fox_Nam_ were predicted to have 1.3x larger HAI (comparable to 1.5x found experimentally) than subjects with no prior infection (**Fig S10A**). Little-to-no antibody ceiling was both measured and predicted for older variants circulating before 2000. For later variants, the model predicted a 2x reduction in fold-change for each 34x increase in pre-vac HAI (while experiments found that only a 10x increase is needed, **Fig S10B**). This effect is small, as increasing pre-vac HAI from 10→320 (the range of most pre-vac titers for these viruses) is predicted to blunt fold-change by 2x (while experiments report a 2.8x blunting). Thus, the model recapitulates these phenomena, although with smaller effect sizes.

Given this caveat, we implemented a robust search for pre-vaccination features that distinguish strong vaccine responses (defined as GMT_post_ ≥2.5ꞏGMT_pre_ against the full variant panel) from weak responses (GMT_post_ ≤1.2ꞏGMT_pre_, **Fig 5A**), while accounting for potential prediction error and experimental error. Unlike the prior sections analyzing absolute HAI, the following analysis uses fold-change (FC) pre-to-post vaccination, often deemed to be an equally important metric of the response.

**Figure 5.**
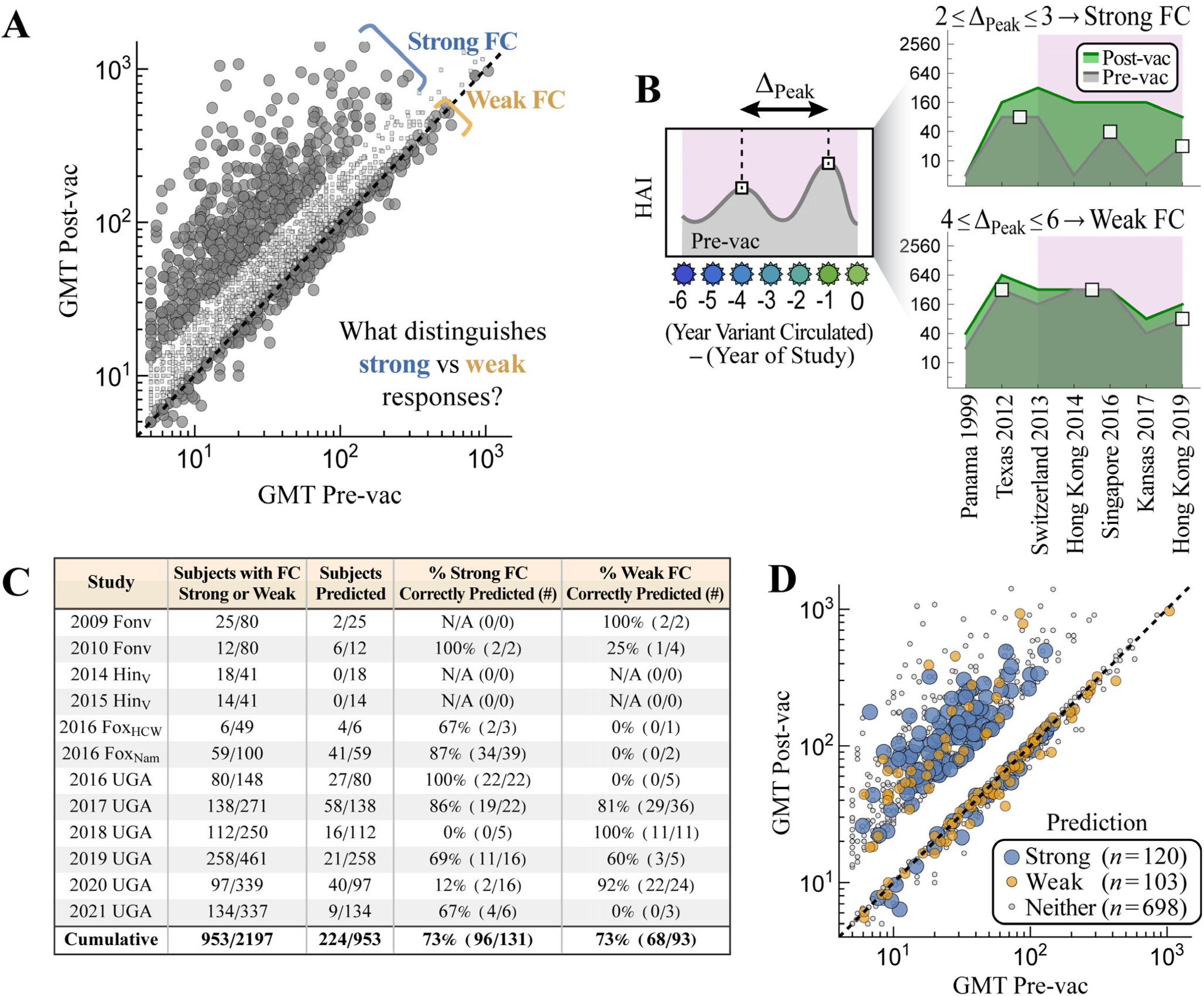
Time between HAI peaks of recent influenza variants distinguishes strong from weak fold-change responses. (A) Vaccine responses were defined as strong fold-change (FC≡post-vac GMT/pre-vac GMT≥2.5) or weak fold-change (FC≤1.1) across all variants; intermediate FC was not considered. (B) Δ_Peak_ was defined as the years between the two most recent HAI peaks (white squares) for variants circulating 0-6 years prior to the study (pink region, **Methods**). Subjects were classified as strong if 2≤Δ_Peak_≤3 and weak if 4≤Δ_Peak_≤6; subjects with longer Δ_Peak_ or ≤2 peaks within this time period were not classified. Data shown for IDs 208 [top right] and 50C [bottom right] from the 2019 UGA dataset. (C) Classification accuracy across all studies. (D) Classifications mapped back onto the pre– and post-vac GMTs for the strong and weak fold-change responses across all studies.

To account for potential model and experimental error, we assessed which subjects not only exhibited either a strong/weak FC response, but where small perturbations (increasing or decreasing any variant’s pre-vac HAI by 2x) were predicted to elicit this strong/weak FC response ≥75% of the time (**Fig S11**). The resulting “robustly strong” and “robustly weak” pre-vac states were further validated by using a support vector machine and nearest neighbor classification system trained on the strong/weak FC responses in each dataset and then used to classify all perturbed states (**Methods**).

The robustly strong HAI responses were noticeably more jagged than the robustly weak responses, especially against recent variants (**Fig S11**). The time Δ_Peak_ between the two most recent pre-vac HAI peaks (for variants circulating within 6 years of each study, **Methods**) correlated with post-vac fold-change. 73% of strong responders had a gap of Δ_Peak_=2-3 years while 73% of weak responses had a larger gap of 4-6 years (**Fig 5B-D**), with a significant decrease in Δ_Peak_ between both classes (*p*<0.001, one-sided permutation test). As a point of comparison, using age≤65 or pre-vac GMT≤80 as an indication of weak-fold change tends to accurately predict strong responses (83-92% accuracy) at the cost of characterizing weak responses (17-37% accuracy, **Fig S12**). In summary, the periodicity of pre-vac HAI against recent variants predicts the fold-change of the post-vaccination response, and such features should be considered in conjunction with features such as age or pre-vaccination titers.

## Discussion

This work developed a computational framework that uses an individual’s pre-vaccination HAI to forecast their response 1-month post-vaccination. Any such model must be able to handle the heterogeneous immune responses across the population, updates to the vaccine composition, and the myriad differences in study designs.

Our approach used: (1) pre-vac HAI against the vaccine strain and (2) pre-vac HAI against historical variants, both of which improved prediction by ∼30% relative to predictions based solely on demographic features such as age. While many studies measure the vaccine strain, data from variants is less common, yet recent work showed that variants can reveal long-term trends in the antibody response^58^ or estimate protection against future variants.^59^

In the context of vaccine responses, measuring variants offers three main advantages. First, prior work has shown that titers from multiple variants can be used to infer exposure history,^60,61^ a feature that is known to affect the antibody response yet is difficult to accurately report. Conceptually, variants break up potential degeneracies in the data: for example, two subjects may have the same pre-vac HAI=20, but in one case it was caused by a minor recent infection while in the other case it was caused by a larger infection several years back. Measuring the response against historical variants would distinguish these two scenarios.

Second, variants provide common features that can be used for prediction, even among studies that administer different vaccines. Even the oldest 1997 Fonville study had ≥4 overlapping variants with every other vaccine study we considered from 1998-2018, letting us test how far forward in time the patterns in early studies continue to hold.

Third, previous work showed that measuring a serum’s pre– or post-vac HAI against as few as 4 variants can predict other variants’ HAI at this same time point (*e.g.*, one pre-vac response can predict another, or that post-vac can predict post-vac).^31^ This demonstrated that at each slice of time, there are conserved patterns between variants that hold across datasets, and in this work we pushed further by using variants to predict the pre-to-post vaccine response.

In comparison to variant pre-vac HAI, a person’s age, sex, BMI, date of vaccination, vaccine dose, or geographics location led to small improvements in prediction accuracy. This lack of predictive power is especially surprising for age, which many studies have shown affects the influenza antibody response.^23,40,42,62^ One explanation is that these age effects may be small (as shown for the infection history and antibody ceiling effects), or they may be dataset-specific and change across seasons.

Indeed, children’s responses (2014-2015 Hinojosa_V_) accurately predicted adult responses and vice versa (upper quartile RMSE<4x). Aside from the two oldest studies, the only poorly predicted dataset was 2015 Hinojosa*_U_*, a cohort containing subjects that self-reported as being uninfected and unvaccinated in the past five years.^37^ Notably, even the 2016 Fox_Nam_ cohort comprising adults receiving the first influenza vaccine of their life was well-predicted. Taken together, these results suggest that a lack of recent influenza infections (and possibly young age) can result in fundamentally different antibody responses.

If these results translate into animal studies, they suggest that experiments on naive mice or ferrets may have limited transferability to humans not only because they are a different species, but also because of their nonexistent immune history. This hypothesis can be tested by comparing the vaccine response in humans to that in mice/ferrets with different exposure histories.^63–66^

Since most influenza deaths occur in the elderly (age 65+), antibody responses in this age group are particularly important. In the UGA studies, all elderly individuals ≥65 years old in the UGA studies were offered high-dose Fluzone, and 76%(=259/340) received it (see the supplemental dataset for each person’s vaccine dose). However, we found no stark difference between elderly individuals (age 65+) receiving high dose vaccines and an adjacent age group (55-64) receiving standard dose Fluzone in any season. Notably, several prior studies found that high dose vaccines elicited higher HAI or led to fewer hospitalizations,^67–69^ although some studies found little effect.^70^ In the UGA studies, the high-dose vaccines may have blunted differences in the response across age groups.

Few methods exist to predict the post-vac response from pre-vac data. Antigenic cartography uses naive ferret data to predict the relationship between the HAI of each strain,^4,59^ but it cannot predict the magnitude of the response to distinguish weak from strong responders. Other methods have predicted whether the vaccine strain’s HAI is above or below a threshold rather than predicting absolute titers.^27,71^ The null and linear models introduced here can predict responses from multiple variants, yet both were more prone to large prediction errors.

Our approach used random forests to predict each variant’s post-vac HAI using its pre-vac HAI together with the four most informative other variants from each study (**Methods**). Information about each vaccine was not directly encoded, but rather indirectly learned in a data-driven manner by training on all studies from the past 10 seasons. This crude encoding cannot distinguish between vaccine formulations or even vaccine strain composition, and hence the overall 2.4x prediction error across all studies suggests that either feature has a minor effect on the resulting vaccine response. This holds across all seasons, including cases where the H3N2 vaccine strain stayed the same (2016→2017, 2022→2023) or changed across clades (2018→2019 [Kansas 2017]→2020). Had a different variant been chosen for the 2019 vaccine, we hypothesize that vaccine studies would have looked similar (but with the new vaccine strain replacing Kansas 2017).

Regarding vaccine formulation, although most studies we analyzed administered Fluzone, other inactivated vaccines (Afluria, Fluarix, Vaxigrip) were predicted with uniformly low error. The only notable exception was the live attenuated vaccine FluMist that elicited no antibody response in 84% of individuals (pre-vac HAI ≈ post-vac HAI for all variants), and future work will determine where other vaccine types (*e.g.*, recombinant, adjuvanted) fall along this spectrum. Note that while inactivated vaccines induced a measurable antibody response on average, ∼35% of individuals still exhibited little-to-no response (≤2x fold-change against any variant).

Across the influenza seasons with predictions (2009-2010, 2014-2023), our approach characterizes ∼88% of sera within the roughly 2x error of the HAI assay. Yet the most extreme responses ‒ the very strongest and very weakest ‒ were poorly predicted, likely because the vaccines elicit a small response in most individuals. Given that the vaccine response peaks 1-month post-vac, the 3% of overpredicted sera may represent individuals who had been infected in the month prior to vaccination, edge cases poorly handled by the model, or experimental error. The 9% of underpredicted responses (comparable to the ∼10% annual influenza incidence^72^) may similarly represent breakthrough infections that magnified the HAI response or a shortcoming of the model.

Yet a model need not perfectly predict all data to be useful. By identifying states that were robustly strong or robustly weak (even after a 2x perturbation to any pre-vac titer), strong responses were found to be associated with jagged HAI landscapes with more frequent peaks. A cross-sectional study recently found long-term periodicity in the influenza response, yet it is unclear what mechanisms underlie such behavior.^58^ We observed short-term periodicity, and we speculate that the oscillatory HAIs from robustly strong responders imply that they elicit antibodies targeting variant-specific epitopes, and hence they are prone to respond strongly to drifted vaccine strains. Conversely, robustly weak responders may target more conserved epitopes that can inhibit and attenuate future vaccine strains with these same conserved sites. Our work is limited in so far as it points out but does not directly explore the underlying biology.

Each vaccine study requires substantial time and effort, but the sera collected have utility beyond quantifying the vaccine response in a single season. For example, existing sera tested against next year’s vaccine variants (as done in 2009/2010 Fonville, 2016 Fox_Nam/HCW_, 2019/2020/2021 UGA) informs future vaccine responses. Alternatively, vaccine responses from early in the season or from studies from the southern hemisphere (whose influenza season is offset by 6 months) could augment predictions in the northern hemisphere.

Predicting vaccine responses at the start of a season sets the stage to design better vaccine strategies. For example, individuals who would exhibit a weak vaccine response with one influenza formulation may benefit from receiving a different formulation. If we could estimate the durability of each person’s response, individuals whose HAI rapidly decays could get booster doses. Next generation vaccines under development will enable more complex vaccine strategies, and while a universal one-size-fits-all vaccination scheme may ultimately result, it is also worth considering vaccine recommendation schemes that better utilize the tools at hand.

## Methods

### Datasets analyzed

Vaccine studies are described in the following manuscripts: Fonville,^4^ Hinojosa,^37^ Fox,^38^ UGA,^39^ Crotty [this work]. Vaccine strains are listed in **Fig S1**; in 14/16 studies the vaccine strain was included in the variant panel, but in the two cases the closest variant was used instead (2009 Fonville: A/Perth/27/2007 [ΔAA_Epitope_=1 substitution in the HA head compared to A/Brisbane/10/2007], 2021 UGA: A/Tasmania/503/2020 [ΔAA_Epitope_=2 substitutions compared to A/Cambodia/e0826360/2020]).

The explicit time points measured in each study can subtly differ. 2016 Fox_Nam/HCW_ and 2016/2017 UGA measured the response 21 days post-vac, whereas all other vaccine studies measured the day 28 post-vac response. In 2020-2021 UGA, measurements were given as a spectrum of more precise dates (*i.e.*, participants were asked to return 28 days post-vaccination, but the exact date of the post-vac visit was recorded. This exact date ranged from 21-71 days, and a similar spread is expected in all other studies). Such subtleties in timing were ignored in this analysis, with all post-vaccination time points treated as “1-month post-vaccination,” which simplifies time from a continuous variable to a Boolean variable.

### Variants

The full list of H3N2 variants measured in each dataset are given in **Fig S3C**. The four new vaccine studies conducted in this work measured HAI titers against A/Hong Kong/4801/2014 (except 2023 UGA), A/Singapore/INFIMH-160019/2016, A/Kansas/14/2017, A/South Australia/34/2019, A/Hong Kong/2671/2019, A/Tasmania/503/2020, and A/Darwin/9/2021. The majority of viruses were propagated in embryonated chicken eggs in the lab of Dr. Ted Ross for all four new vaccine studies. The only exceptions were A/Darwin/9/2021 and A/Kansas/14/2017 in the 2023 Crotty_Afluria/FluMist_ studies, which due to limited reagents were instead supplied by Dr. Florian Krammer.

### Vaccine Study Participants

25 participants were recruited for each of the four new influenza vaccine studies presented in this work (2022 UGA, 2023 UGA, 2023 Crotty_Afluria_, and 2023 Crotty_FluMist_). These studies administered different formulations of the influenza vaccine from that season.

The 2022 UGA study administered the 2022-23 vaccine comprising H1N1 A/Victoria/2570/2019, H3N2 A/Darwin/9/2021, B/Austria/1359417/2021 (B/Victoria lineage), and B/Phuket/3073/2013 (B/Yamagata lineage). As in prior UGA studies, participants less than 65 years old were given standard-dose [15 mg/component] Fluzone Quadrivalent (Sanofi Pasteur). Participants aged 65 or older were offered the high-dose [60 mg/component] Fluzone Quadrivalent (Sanofi Pasteur), and 3/5 of participants in this age group opted for the high-dose vaccine (see the supplemental dataset for information on vaccine dose).

All three 2023 studies administered the 2023-24 vaccine composed of H1N1 A/Victoria/4897/2022 and the same H3N2, B Victoria, and B Yamagata strains as the 2022-23 vaccine. In the 2023 UGA study, participants under 65 years old were given standard-dose Fluzone Quadrivalent (Sanofi Pasteur) while those 65 and older were offered the high-dose version, with 9/10 participants in this age group opting for the high-dose vaccine. All participants in the 2023 Crotty_Afluria_ study received the 2023-24 formulation of Afluria Quadrivalent (Seqirus) while those in 2023 Crotty_FluMist_ received FluMist Quadrivalent (AstraZeneca).

In all studies, sera were collected pre-vaccination (day 0) and one-month post-vaccination (day 28) in the fall of their respective year. Participants for the 2022-2023 UGA studies were recruited from medical facilities near Athens GA while those in the Crotty studies were recruited from La Jolla CA. The SST tube of one participant in 2023 Crotty_Afluria_ clotted during the one-month blood draw resulting in *n*=24 post-vaccination samples, and the unpaired pre-vaccination sample was dropped from our analysis.

The UGA vaccine studies were approved by the Western Institutional Review Board and the University of Georgia Review Board. The Crotty studies were approved by the La Jolla Institute for Immunology Review Board (IRB#: VD-271).

### Analyzing HAI Titers

The hemagglutination inhibition (HAI) assay quantifies how potently an antibody or serum inhibits the ability of a virus to bind red blood cells. The value (or titer) for each antibody-virus pair corresponds to the maximum dilution at which an antibody inhibits this interaction, so that larger values represent a more potent antibody. This assay is traditionally done using a series of 2-fold dilutions, so that the HAI titers can equal 10, 20, 40… Plots with too many overlapping points were jittered for clarity (**Figs 2B, 3B, 3F, 4B**).

As in previous studies, all analysis was done on log_10_(HAI titers) because experimental measurements span orders of magnitude, and taking the logarithm prevents biasing the predictions toward the largest values while also accounting for the declining marginal protection from increasing titers.^10^ For example, the dashed line in **Fig 1B** shows that the vaccine strain’s geometric mean titers post-vac equal 95; using the arithmetic mean would yield mean post-vac titers of 230.

Prediction error was quantified in unlogged units so that it can be readily compared to the measured values. RMSEs were computed by first taking the root-mean-squared error *σ* of the log_10_(HAI titers) and then present the un-logged value that is exponentiated by 10 (*i.e.*, *σ*=0.3 for log_10_ titers corresponds to an error of *σ*_Predict_=10^0.3^=2-fold, with “fold” or “x” indicating an un-logged number). Confidence intervals were computed by bootstrapping the log_10_ values.

### Intrinsic Noise of the HAI Assay

Harvey *et al*. reported HAI from influenza surveillance data, where HAI from the same ferret sera were measured against the most common circulating variants each week.^73^ Thus, their dataset contained ∼700,000 repeat measurements of the same serum-virus pair, and these measurements were consistent with Gaussian error (on a log_2_ scale) with standard deviation *σ*=1 (i.e. 2-fold error). More precisely, 40.0% of their repeat measurements did not change (1x error), 44.7% had 2x error, 12.6% had 4x error, 2.4% had 8x, and 0.2% had 16x error. In comparison, a log-Gaussian error distribution would predict that 38.7% of measurements would not change, 48.3% have 2x error, 11.7% have 4x error, 1.1% have 8x error, and 0.03% have 8x error.

In addition, Fonville *et al*. analyzed HAI measurements from nearly identical sera and found that the inherent error of the assay is log-normally-distributed with standard deviation ≈2-fold.^4^ This is shown by Figure S8B in Fonville *et al*.^4^ (neglecting the stack of not-determined measurements outside the dynamic range of the assay), where 40% of repeats had the same HAI value, 50% had a 2-fold discrepancy, and 10% had a 4-fold discrepancy.

As further corroboration, across the 15 studies examined there were *n*=9 pairs of variants with identical HA sequences that were measured within the same study (points in **Fig S3A** with ΔAA_Epitope_=0 that also all had ΔAA_Total_=0). The RMSE between the HAI of these 9 variants varied from 1.6x-3.3x with a geometric mean of 2.1x.

### HAI Protocol

For the UGA studies, sera were treated with receptor destroying enzyme (RDE) (Denka) to inactivate nonspecific inhibitors. Briefly, three volumes of RDE were added to one volume of sera and incubated overnight at 37°C. The next day, samples were incubated at 56°C for 30-60 minutes, after which 6 volumes of 1x phosphate buffered saline (PBS) were added to each sample, resulting in a final serum dilution of 1:10. For the Crotty studies, 20µL starting volume of serum was treated with 0.5×(starting volume) of 8mg/mL TPCK-trypsin (Sigma-Aldrich) and incubated at 56℃ for 30 minutes. After cooling to room temperature, 3×(starting volume) of 11mM potassium periodate (Sigma-Aldrich) was added and incubated for 15 minutes at room temperature. 3×(starting volume) of 1% glycerol-PBS was then added and incubated for 15 minutes. Lastly, 2.5×(starting volume) of 0.85% PBS was added to all samples.

Serum was diluted in a series of 2-fold serial dilutions in 96-well V-bottom plates (Thermo Fisher for UGA studies, Sarstedt for Crotty studies). An equal volume of influenza virus, adjusted to 8 hemagglutination units (HAU)/50µL diluted in 1x PBS, was added to each well of the plate. For the UGA studies, plates were then covered and allowed to incubate at room temperature for 20 minutes. After incubation, 50µL of a solution consisting of 0.8% turkey red blood cells (Lampire Biologicals) diluted in 1x PBS was added to each well. The plates were then mixed by gentle agitation, covered, and allowed to incubate for another 30 minutes at room temperature. For Crotty studies, sera and virus were incubated for 30 minutes at room temperature. After this incubation, 50µL of 0.5% turkey red blood cells (Lampire Biologicals) or 0.75% guinea pig red blood cells (GPRBCs) in 1x PBS (Lampire Biologicals) were added to all wells and incubated at 4℃ for 45 minutes. GPRBCs were used for H3N2 A/Darwin/9/2021 in the Crotty studies due to lack of viral activity observed with turkey red blood cells.

After incubation with red blood cells the plates were tilted to observe the hemagglutination inhibition. The HAI antibody titer was determined by taking the reciprocal dilution of the last well that contained non-agglutinated red blood cells. For GPRBC plates, the HAI titer was determined as the last dilution with a “halo” of the same size as PBS and virus only wells. Samples with no detectable activity were assigned to half the limit of detection (HAI=5).

To confirm assay consistency between runs, positive controls were included. In the UGA studies, these consisted of sera from previously performed mouse or ferret infections. In the Crotty studies, normal control goat serum (FR-1377) and positive control influenza A(H3) Reference Goat Antiserum (FR-1562, FR-1612, FR-1683, FR-1737, FR-1780, FR-1827) were obtained through the International Reagent Resource, Influenza Division, WHO Collaborating Center for Surveillance, Epidemiology and Control of Influenza, Centers for Disease Control and Prevention, Atlanta, GA, USA. In addition, pre– and 1-month post vaccination serum for the same participants were run on the same plate together with the negative and positive control serum.

### Quantifying Predictive Power of Different Features

A pair of subjects matched across each feature in **Fig 2A** if they met the following conditions:

● *Age*: Pairs have ages ≤10 years apart from one another. (Alternative thresholds of ≤5 and ≤20 years apart led to worse predictions.)
● *Sex*: Pairs have the same sex (male or female).
● *BMI*: Pairs have a BMI ≤2 of one another. (A threshold of ≤5 led to worse predictions.)
● *Date of vaccination* [1=Jan 1, 365=Dec 31]: Pairs have a date of vaccination ≤14 days apart. (A threshold of ≤21 or ≤28 days made no difference.)
● *Vaccine dose*: Pairs receive the same standard-dose or high-dose vaccine. (The latter is only available [but not mandatory] when age≥65 in the UGA studies.)
● *Location*: Pairs exactly matched along the geographic location of the study (all Fonville studies or 2016 Fox_HCW_ → Australia, 2016 Fox_Nam_ → Ha Nam, all Hinojosa studies → Wisconsin, all UGA studies → Georgia).
● *Pre-vac HAI* (Vac_Pre_): Pairs exactly match in their pre-vaccination HAI titer against the respective vaccine strain in their study.
● *Variant HAI*: Pairs must be measured against ≥5 overlapping viruses (excluding their vaccine strains), and there must be a correlation>0.9 between the log_10_(HAI titers) of these overlapping viruses.

When assessing what dataset features affect the vaccine response (**Fig S4**), predictions were considered both forward and backward in time (*e.g.*, 2016 UGA can predict 2017 UGA and vice versa), since there are cases where predictions appear accurate in one direction but the reverse predictions reveal a dissimilarity. After excluding the 1997/1998 Fonville studies and 2015 Hinojosa*_U_*, all subsequent predictions were predictions forward in time.

### Predicting the HAI Response 1-Month Post-Vaccination

The random forest framework adapted our previous approach,^31^ with each decision tree constrained to use pre-vaccination data as input and post-vaccination data as output. In constructing the random forests, variants *V*_1_-*V*_5_ are chosen with replacement, so that variants can appear multiple times. At least one variant is always set to *V*_0_, since unsurprisingly, the variant-of-interest’s HAI at time 0 was highly informative of its HAI 1-month post-vac.

Row-centering was found to be crucial to accurately predict across datasets. Row-centering means that if *t*_0_-*t*_5_ represent the log_10_(titers) of *V*_0_-*V*_5_, with mean titer *t*_avg_, then each decision tree will take (*t*_1_-*t*_avg_, *t*_2_-*t*_avg_, *t*_3_-*t*_avg_, *t*_4_-*t*_avg_, *t*_5_-*t*_avg_) as input to predict *t*_0_-*t*_avg_. The value *t*_avg_ (which will be different for each serum) is then added to this prediction to undo the row-centering. Note that row-centering does not throw away any information. Similarly, this approach does not normalize or threshold the HAI titers in any way. Instead, the exact post-vaccination titers were predicted for each variant.

When using multiple training sets, predictions from each study were equally weighed by taking their geometric mean (or the arithmetic mean of log_10_[HAI titers]). Unequal weights (for predictions from 1, 2, 3… prior seasons) were assessed through nonlinear fitting, yet we found that the optimal weights were essentially flat and hence opted for equal weighing.

The final post-vac predictions were given as the maximum of the pre-vac HAI and the model predictions. Although there are few cases where model predictions fell below the pre-vac titers, this biologically-realistic constraint slightly improved model predictions.

### Classifying Robustly Strong and Robustly Weak Responses

In addition to our random forest (regression) model, we also trained two classification models using either nearest neighbors or a support vector machine. Both latter models were trained separately on each dataset, taking in every subject’s pre-vac HAI against all variants and a classification of “strong” if post-vac GMT/pre-vac GMT≥2.5 (which requires a substantial post-vac response, especially since HAI from older variants stays nearly constant post-vac) and “weak” if post-vac GMT/pre-vac GMT≤1.1 (a null response accounting for error). Subjects with an intermediate fold-change were not used. The resulting models were then applied to each subject’s pre-vac HAIs as well as a perturbed version of these HAIs (where each titer was increased or decreased by 2x), and cases where ≥75% of states were strong/weak were denoted as robustly strong/weak.

We determined the peaks in HAI data by first sorting viruses by their year of circulation. H3N2 A/South Australia/34/2019 (the 2019 southern hemisphere strain) was removed from 2019-2021 UGA since it interfered with H3N2 A/Hong Kong/2671/2019 (the 2020 northern hemisphere strain) and allowed peaks to happen in 2019 and 2020 which decreased classification accuracy. Peaks were then determined after applying a Gaussian blur with scale *σ*=1 to account for error. In case of a plateau where multiple consecutive variants had an equally large HAI, the peak was set at the position of the earliest variant.

The two most recent peaks between (year of study)‒6 and (year of study) were used to compute Δ_Peak_; if two peaks did not exist in this time frame then no classification was made. Attempts to use values of Δ_Peak_>6 to classify lead to worse accuracy, likely because the variant panels lose resolution beyond this time interval and hence many peaks are artificially missed.

## Data Availability

The data and code used in this work is contained in the following GitHub repository: https://github.com/TalEinav/PredictFluVaccine

Both the raw data and random forest predictions are provided in a CSV file. We also provide a supplementary Mathematica notebook that implements our prediction framework and reproduces all figures from this manuscript.

## Author Contributions

**Conceptualization:** Tal Einav

**Data acquisition** Hannah Stacey, Michael A. Carlock, James D. Allen, Hannah B. Hanley, Tal Einav

**Formal analysis:** Tal Einav

**Writing – original draft:** Tal Einav

**Writing – review & editing:** Hannah Stacey, Michael A. Carlock, James D. Allen, Shane Crotty, Ted M. Ross, Tal Einav

## Acknowledgements

We especially thank the experimental groups who shared their data, and we hope this paper will inspire other groups to integrate their datasets for everyone’s benefit. We always welcome pointers to new datasets. We further acknowledge the LJI clinical core, UGA influenza vaccine team, UGA Clinical and Translational Research Unit (CTRU), Brad Phillips, and Kimberly Schmitz. Certain influenza A and B viruses were obtained through the Prevention and Control of Influenza, Centers for Disease Control and Prevention, Atlanta, GA, USA and the International Reagent Resource.

This research was supported by the University of California San Diego PREPARE Institute and by the Bodman family (TE), National Center for Advancing Translational Sciences of the NIH (CTRU, Award #UL1TR002378), Collaborative Influenza Vaccine Innovations Centers (CIVICs) of NIAID (Contract #75N93019C00052), University of Georgia, Cleveland Clinic, and by the Georgia Research Alliance (TMR, Georgia Eminent Scholar GRA-001).

## Figures and Tables

**Table S1.** Percent of all participants vaccinated two seasons in a row with consistently strong (HAI≥80) or consistently weak (HAI≤40) responses against the vaccine strain. The right-most column represents the total number of differences in either HA epitopes A-E (ΔAA_Epitope_) or the entire HA (ΔAA_Total_) between the H3N2 vaccine strain’s hemagglutinin (HA) between both seasons. Rows where >80% of subjects showed consistent responses are shaded in gray. The H3N2 vaccine strains are given in **Fig S1A**; note that the H3N2 vaccine strain did not change between the 2016-2017 or 2022-2023 seasons and minimally changed in 2017-2018. The final two rows are based on the 2022/2023 UGA vaccine studies introduced in this work and have far fewer subjects.

| Current → Next Season | % Subjects Strong/Strong or Weak/Weak in Both Seasons (*n*) | $\Delta AA_{\text{Epitope}}$ across vaccine strains (amino acids) | $\Delta AA_{\text{Total}}$ across vaccine strains (amino acids) |
| --- | --- | --- | --- |
| 2016 → 2017 | 83 (*n*=102) | 0 | 0 |
| 2017 → 2018 | 88 (*n*=149) | 3 | 6 |
| 2018 → 2019 | 73 (*n*=161) | 10 | 16 |
| 2019 → 2020 | 64 (*n*=316) | 11 | 18 |
| 2020 → 2021 | 74 (*n*=252) | 11 | 16 |
| 2021 → 2022 | 84 (*n*=19) | 8 | 10 |
| 2022 → 2023 | 100 (*n*=3) | 0 | 0 |

**Figure S1.**
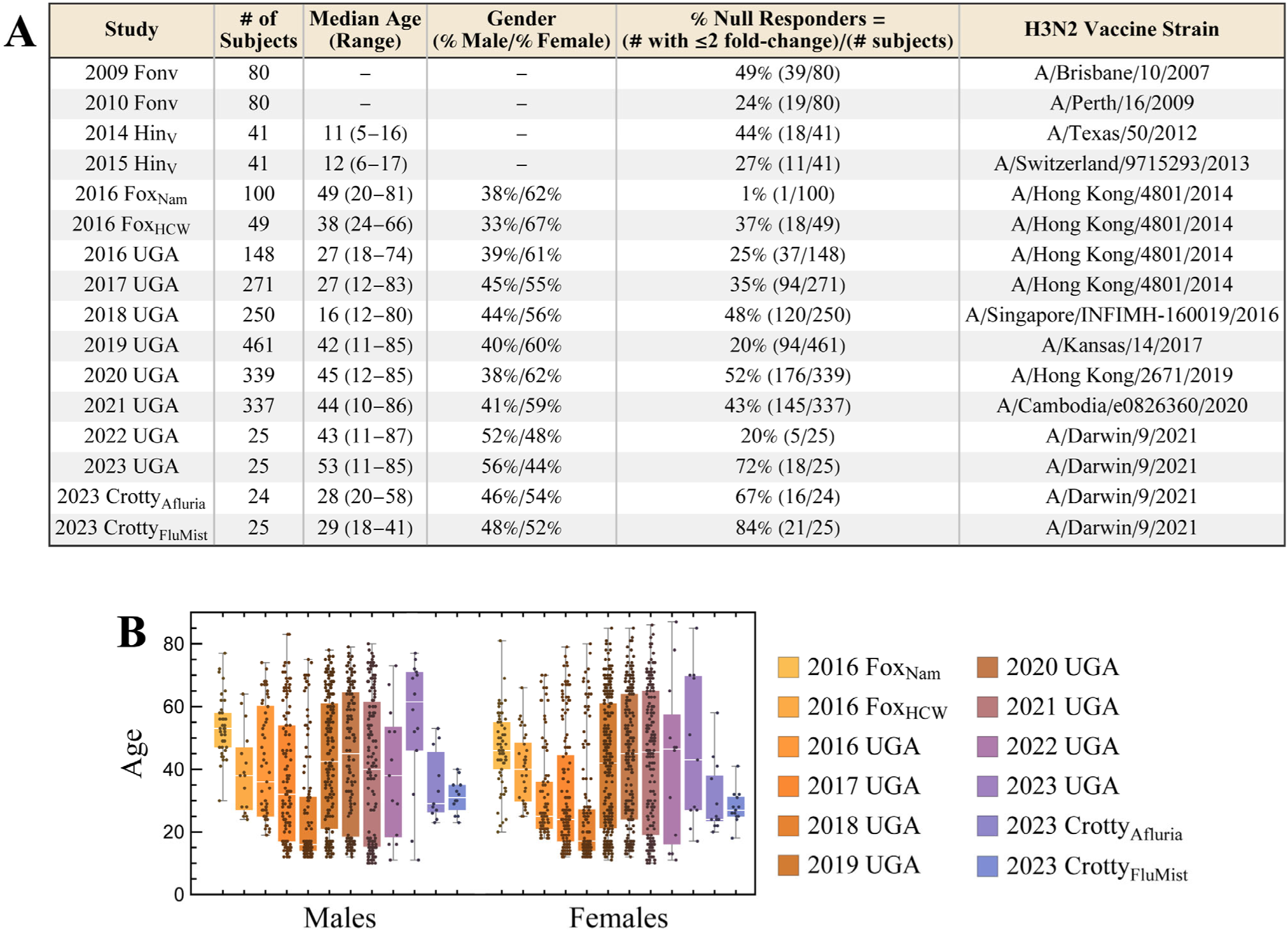
Demographics and null responders in the vaccine studies used to predict responses. (A) Null responders are defined as participants whose response 1-month post-vac differed by ≤2-fold across all variants measured. The final four entries represent the new studies introduced in this work; each study enrolled 25 participants, although one participant in the 2023 Crotty_Afluria_ study did not have post-vac measurements because their serum tube clotted during the blood draw. Dashed represent features that were not reported. (B) Age distribution for all males and females (when measured) in each study.

**Figure S2.**
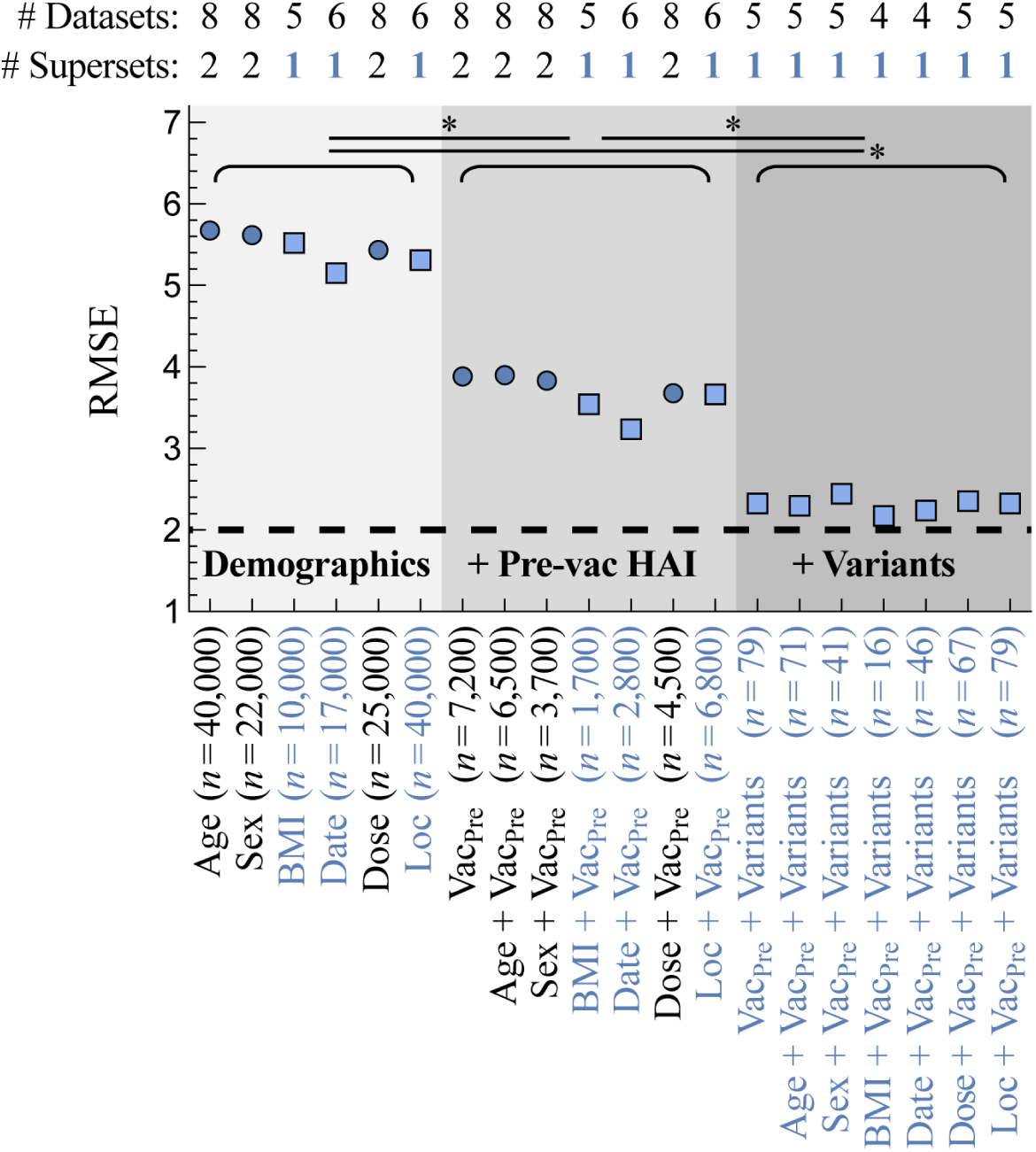
Feature importance when predicting vaccine responses in 65-year-olds or older. Analysis from **Fig 2A** carried out exclusively on subjects with age≥65 (such subjects were only present in the UGA or Fox supersets). For all pairs of subjects vaccinated in different seasons that match across a feature set [*x*-axis], their post-vac HAI is compared against their respective vaccine strains. Features include body mass index (BMI), the date of vaccination (Date), standard vs high dosage (Dose), geographic location (Loc), pre-vac HAI against the vaccine strain (Vac_Pre_), and pre-vac HAI against other variants (**Methods**). Root-mean-squared error (RMSE, *y*-axis) is shown for each feature set, assessed over *n* pairs of subjects. 95% CI are smaller than the plot markers. Blue circles indicate that pairs of matching individuals can be found across multiple studies; teal squares indicate that matched pairs may have artificially small RMSE because they came from a single superset of studies (UGA or Fox; all cases of 1 superset came from UGA studies). The dashed line represents the intrinsic 2-fold error of the HAI assay.^4^ \**p*<0.001 between any feature sets with different gray shading (*e.g.,* Age vs Vac_Pre_ or Date vs Vac_Pre_+Variants) using a one-sided permutation test.

**Figure S3.**
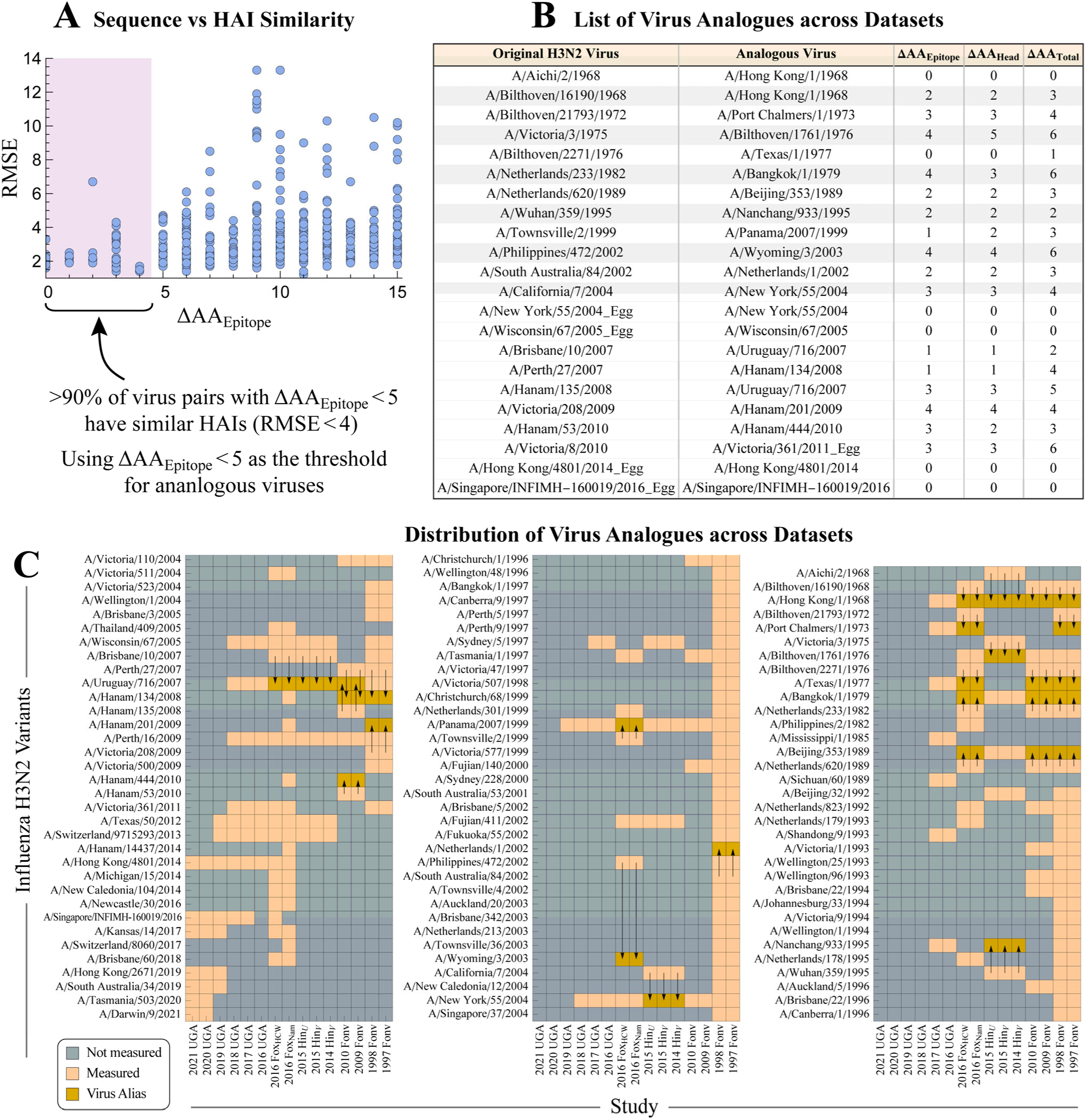
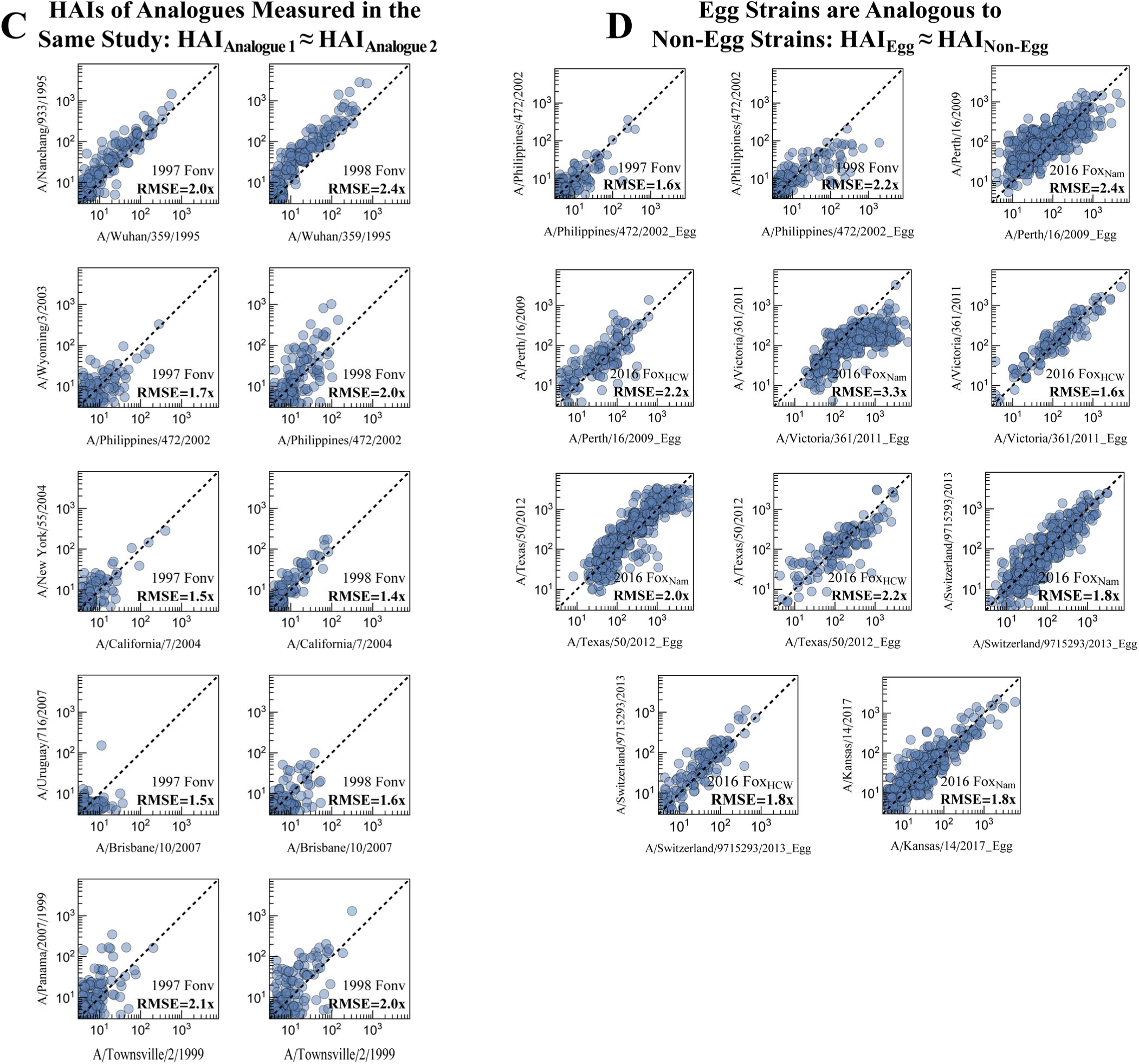
Equating analogous viruses across studies. (A) Determining HAI similarity as a function of amino acid mutations across H3N2 epitopes A-E (ΔAA_Epitope_). Within each dataset, for all pairs of viruses with a given ΔAA_Epitope_ [*x*-axis], we computed the root-mean-squared error between their log(HAI) [*y*-axis]. Virus sequences were taken from each study when available and otherwise from GISAID consensus sequences. (B) Using the threshold ΔAA_Epitope_<5 to equate viruses across studies; viruses were equated whenever it increased the number of predictions (by creating more squares in the same row in Panel C). Differences in the HA amino acid sequence are shown considering H3N2 epitopes A-E [ΔAA_Epitope_], the full HA head [ΔAA_Head_], or the full HA head+stem [ΔAA_Total_] between the original variant and its analogue. Most (but not all) epitopes are within the HA head, so ΔAA_Epitope_≤ΔAA_Head_ is generally true. Both ΔAA_Epitope_≤ΔAA_Total_ and ΔAA_Epitope_≤ΔAA_Head_ always hold. (C) List of variants measured in each study, with arrows representing a shift to an analogous strain to increase virus overlap. All egg– and cell-passaged strains were equated, since in every study when both were measured their HAI titers were approximately identical [Panel D]. (D) HAI similarity directly assessed for cases where two virus analogues were measured in the same study. HAIs from analogue 1 (*x*-axis) and analogue 2 (*y*-axis) across all sera and all time points are shown, with most points lying along the diagonal leading to RMSE≲2x. (E) In the vast majority of cases where an egg and non-egg strain were measured in the same study, the HAI of both was nearly identical with an RMSE≲2x.

**Figure S4.**
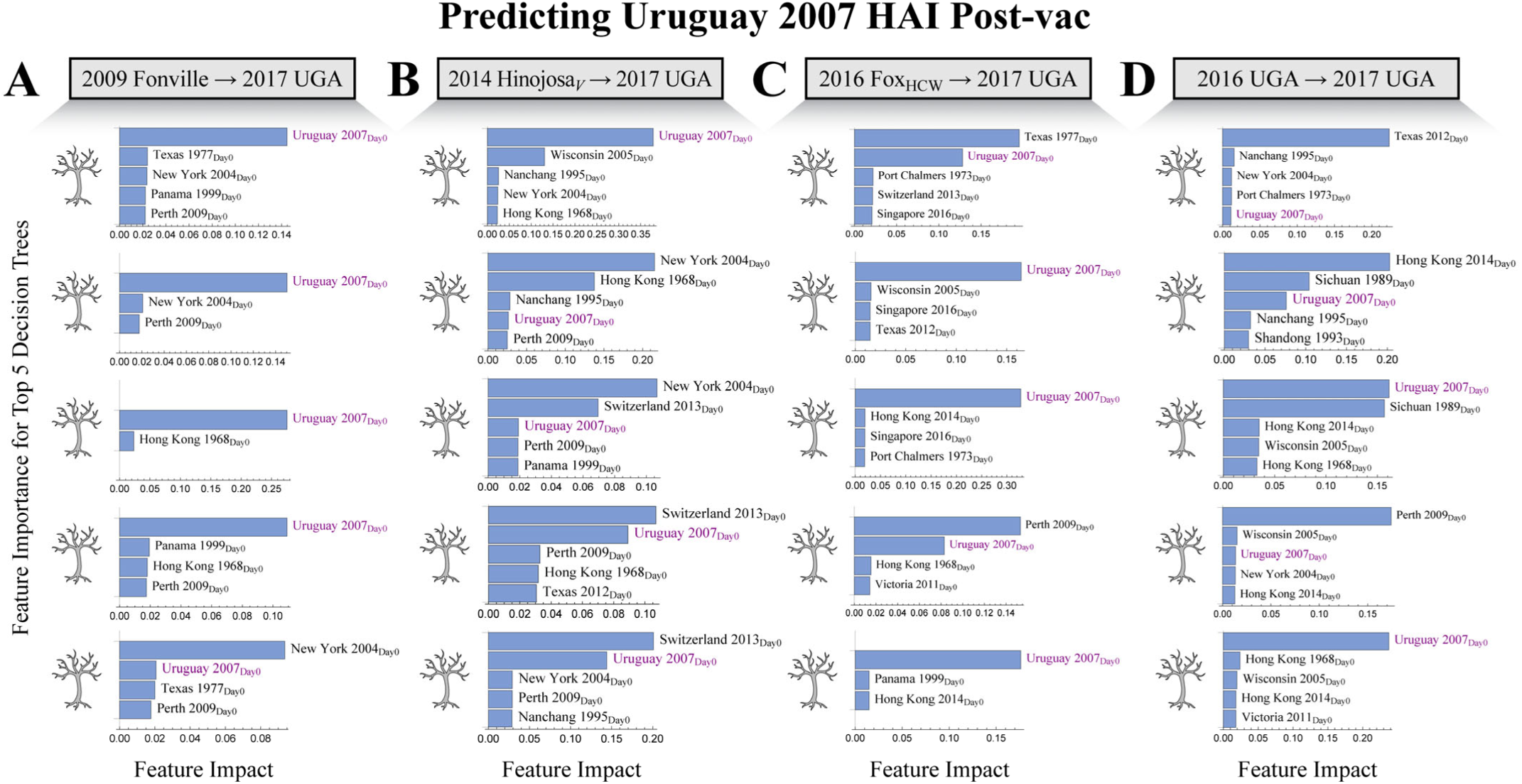
Example predictions for H3N2 A/Uruguay/716/2007 in 2017 UGA. Feature importance when predicting Uruguay 2007’s post-vac HAI using (A) 2009 Fonville, (B) 2014 Hinojosa_V_, (C) 2016 Fox_HCW_, or (D) 2016 UGA. In each case, decision trees were trained using Day 0 HAI from different variants to predict Uruguay 2007’s HAI 1-month post-vaccination. The average predictions from the top 5 decision trees (with the lowest internal RMSE) were used to predict Uruguay 2007’s HAI in 2017 UGA. Plots show the absolute value of SHAP feature importance.

**Figure S5.**
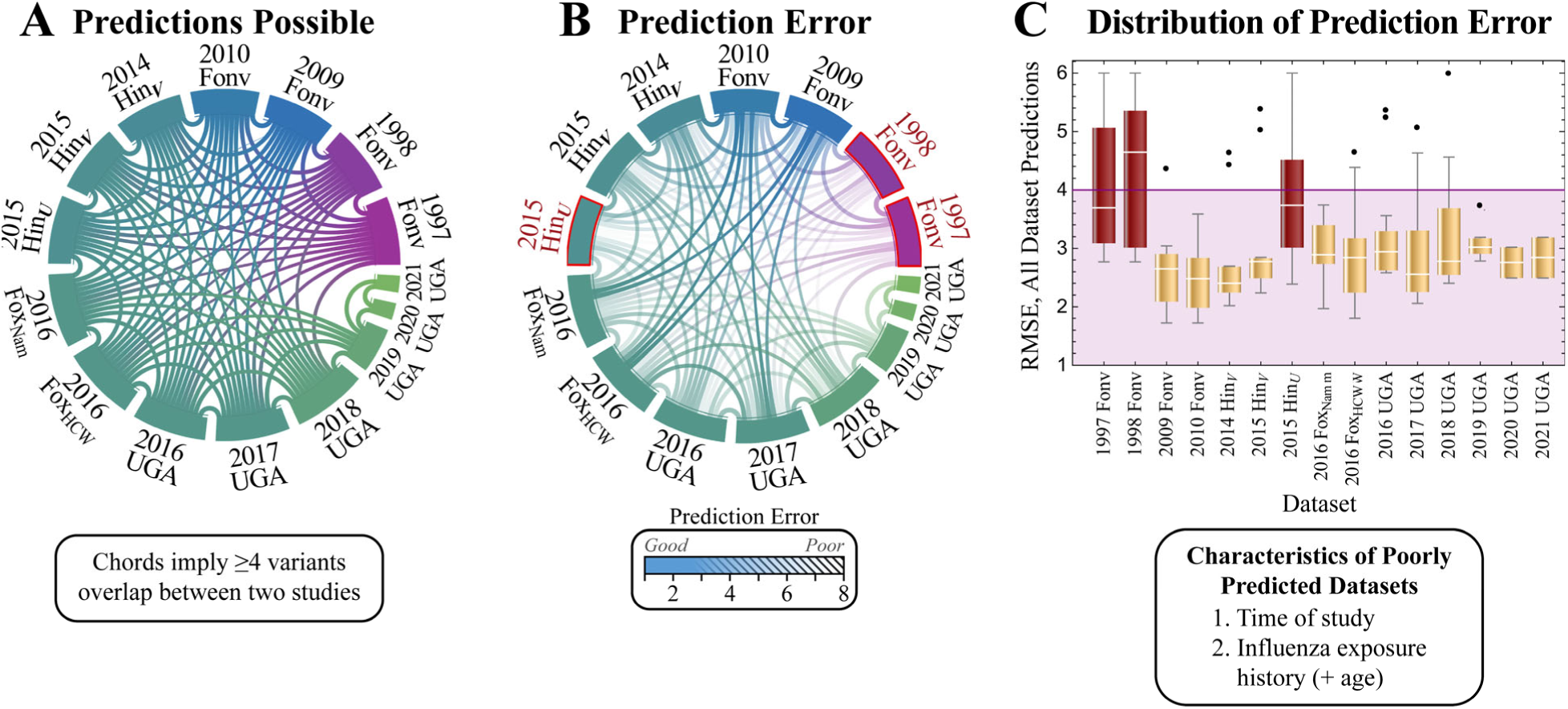
Accuracy of pairwise predictions between vaccine studies. (A) Predictions between two studies were carried out when ≥4 variants were measured in both studies, as indicated by each chord. For clarity, studies are colored from purple-to-green based on the year they were carried out. (B) The error from each pairwise prediction in both directions (average of Study *X* predicting Study *Y* and Study *Y* predicting Study *X*) is shown by each chord’s opacity. Lower opacity represents poorer predictions. (C) Distribution of all predictions to and from each study. The three studies boxed in red (Panel B) or shown in red boxes (Panel C) have an upper quartile RMSE>4x (above the purple region), indicating that some features of their study design led to fundamentally different vaccine responses. The characteristics of these three studies are shown below Panel C.

**Figure S6.**
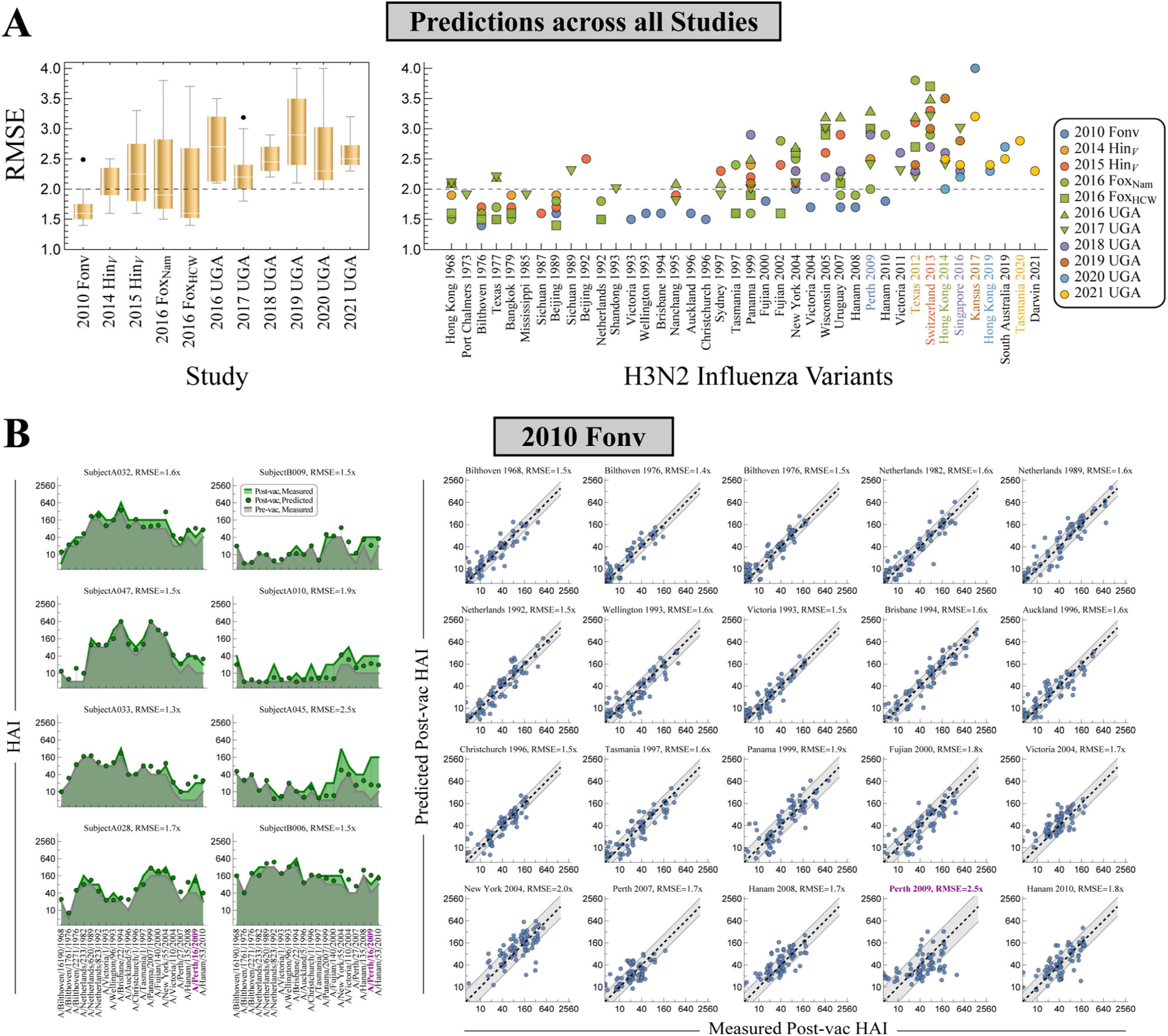
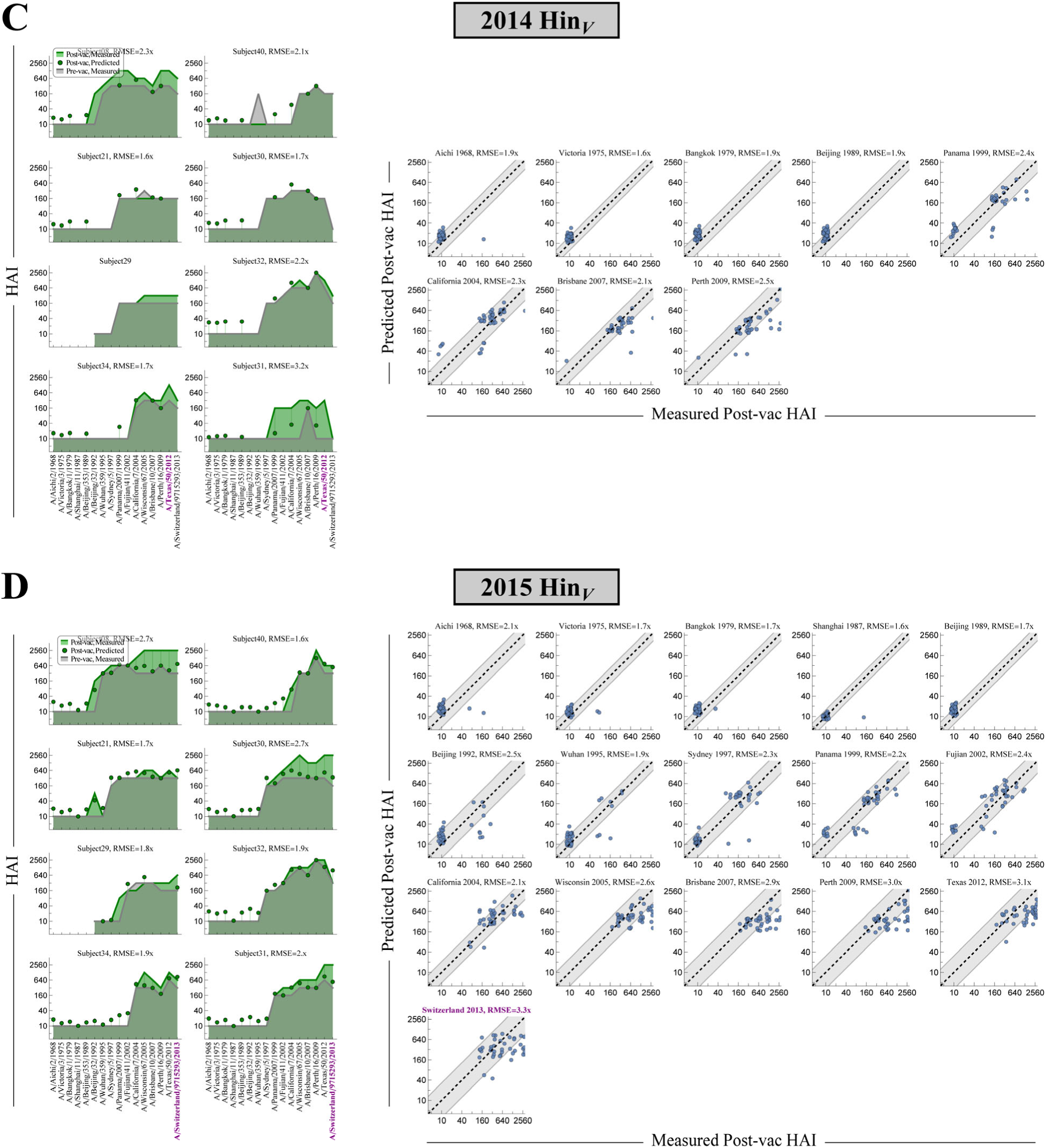
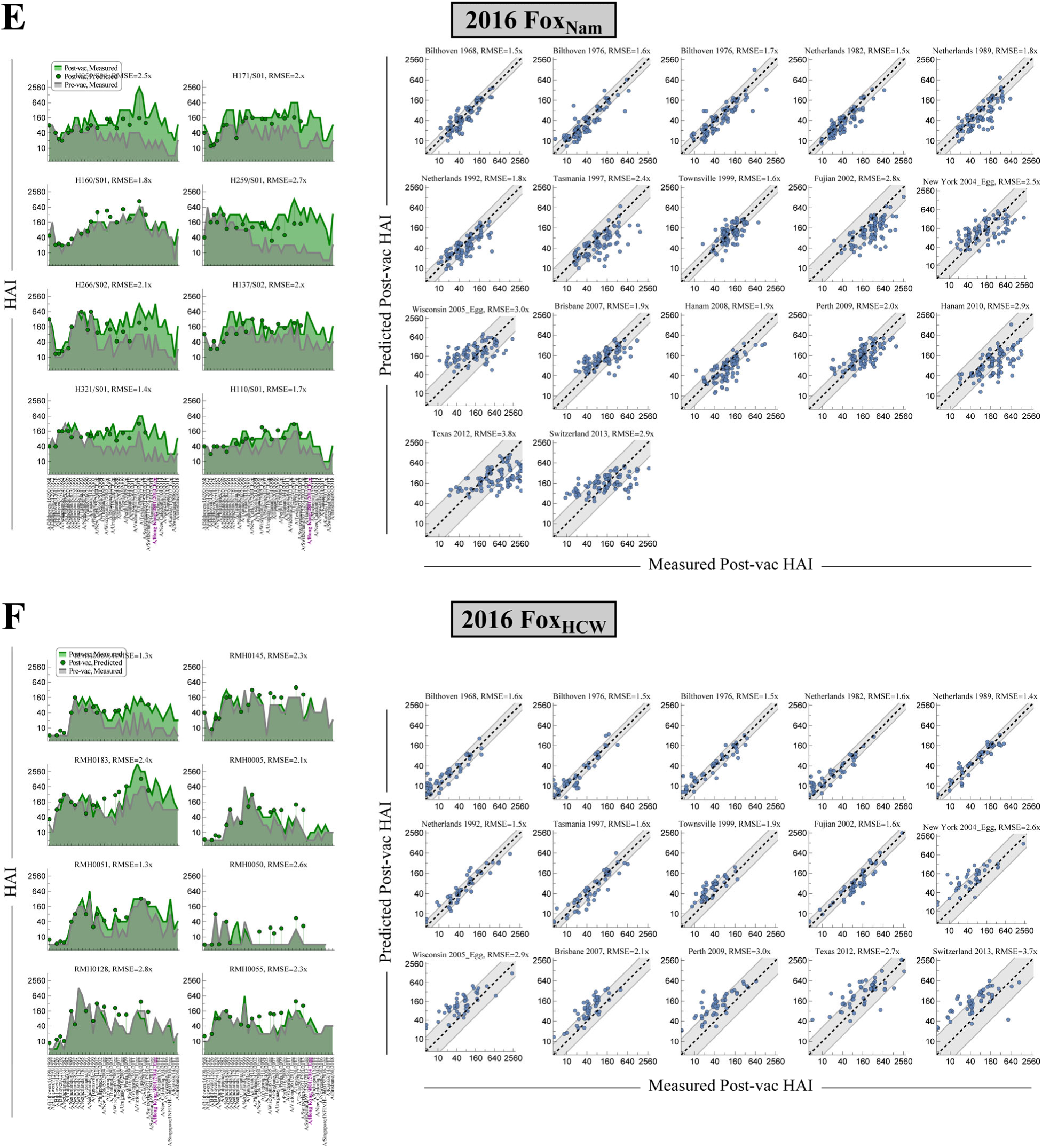
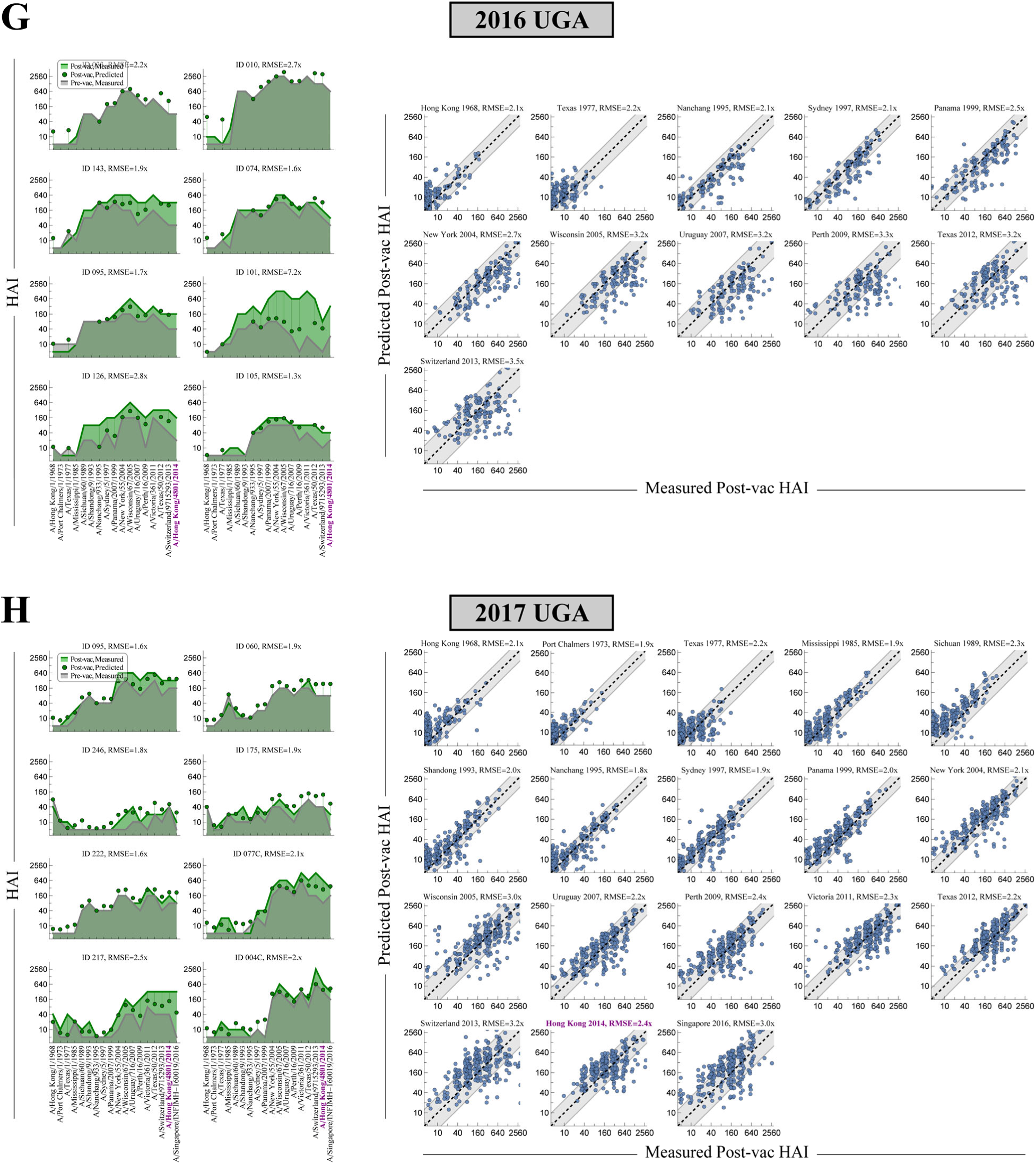
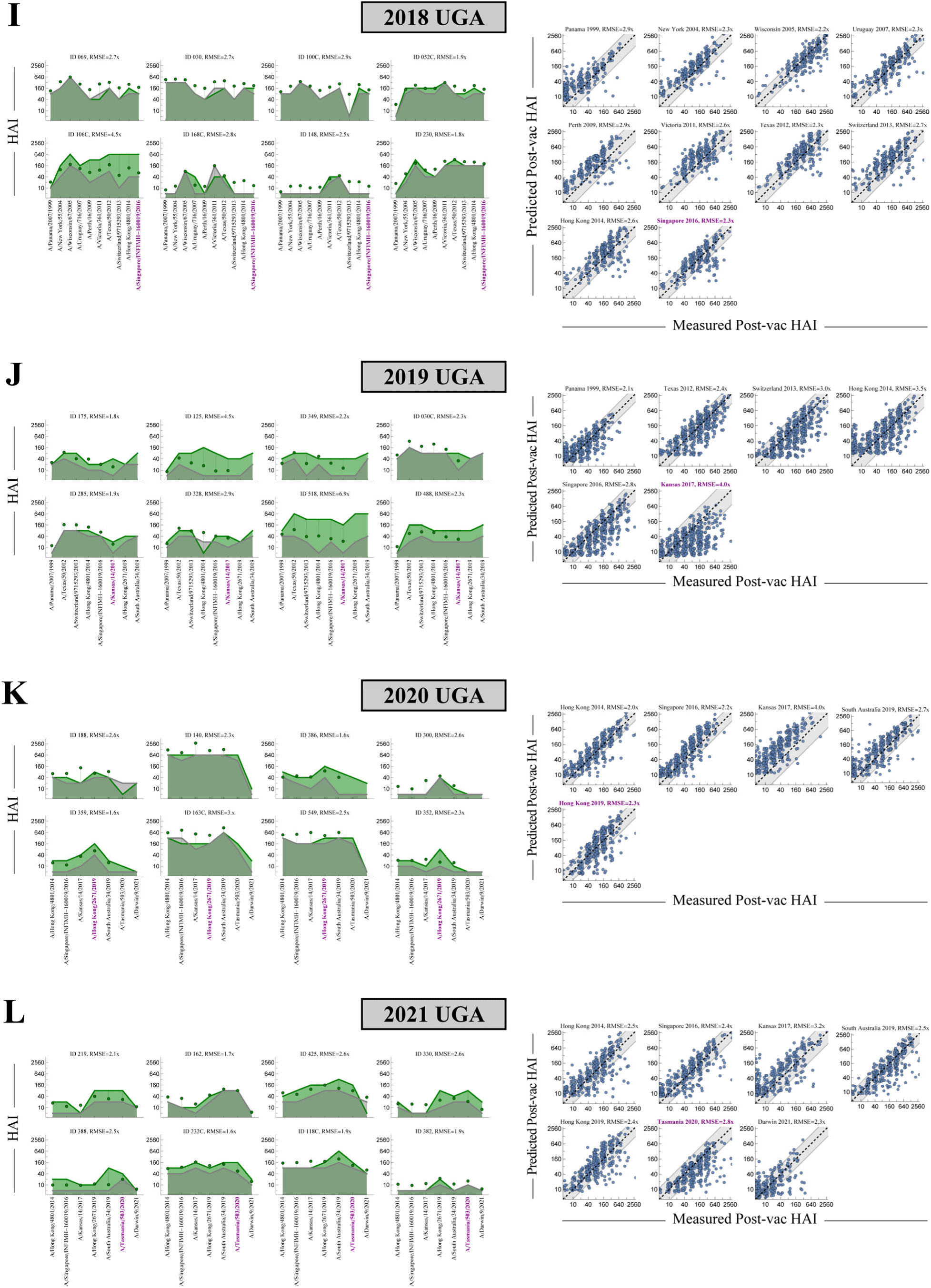
Summary of predictions across all datasets. (A) Distribution of prediction errors across each dataset [*left*] or each variant [*right*, colored variants on *x*-axis denote the vaccine strains]. (B-L) Examples of post-vac predictions for eight randomly chosen subjects [*left*] and up to 20 variants [*right*]. The vaccine strain is highlighted in purple in each study. Some variants may not have predictions if they were not measured in any previous datasets.

**Figure S7.**
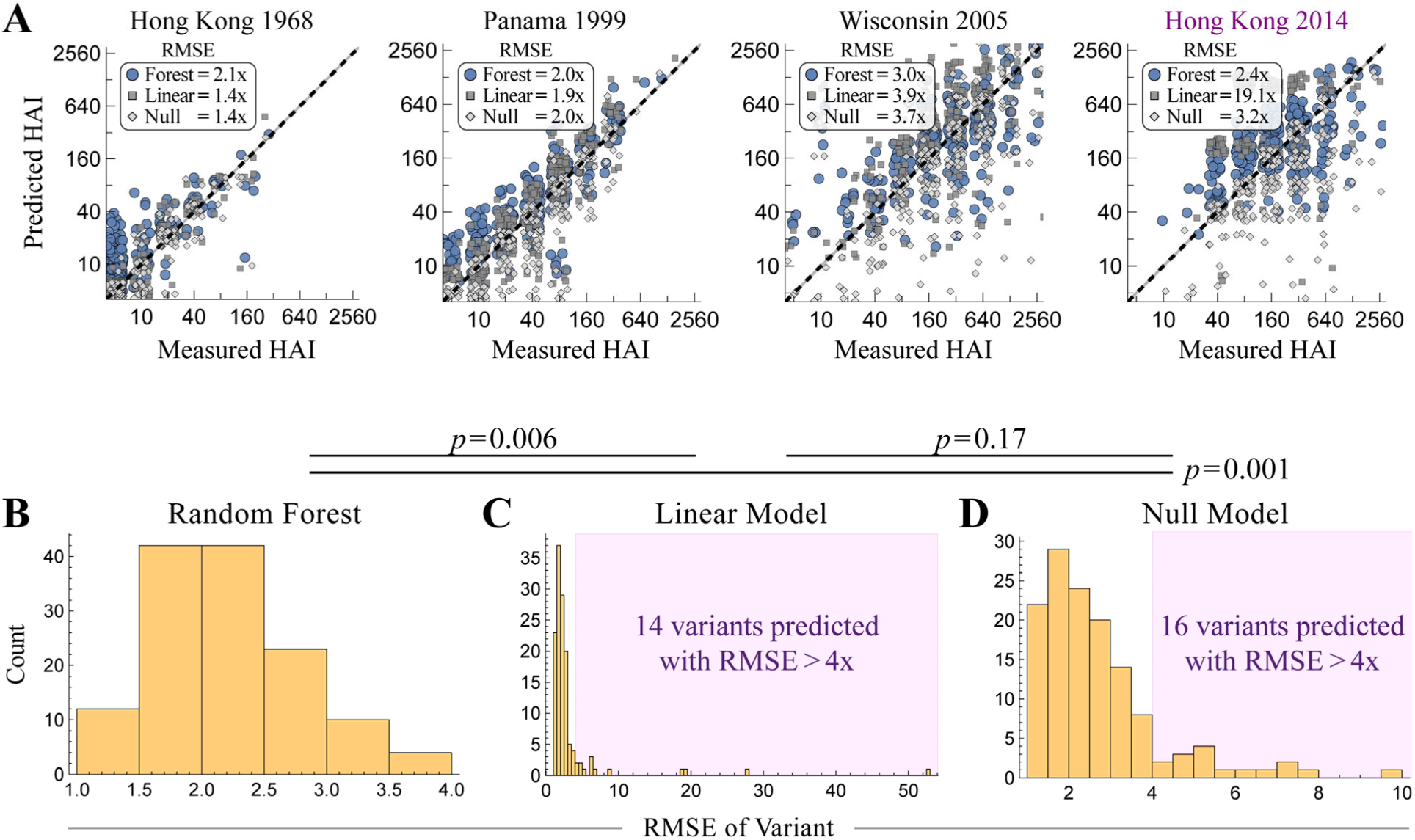
Comparing modeling approaches. (A) Predicted versus measured post-vac HAI of four variants in 2017 UGA, showing the random forest predictions (blue, as in **Fig 3F**) overlaid with the linear model (gray squares) and null model (gray diamonds). (B-D) The distribution of RMSEs for all viruses predicted in each study. (B) The random forest approach described in Fig 3. (C) A linear model predicting each variant *V* by training on all (HAI_V,pre_, HAI_V,post_) titers from prior datasets. (D) A null model assuming that each variant’s post-vac HAI equals its pre-vac HAI. The number of variants predicted with RMSE>4x is emphasized in each panel. *P* values are calculated using a 1-sided permutation test.

**Figure S8.**
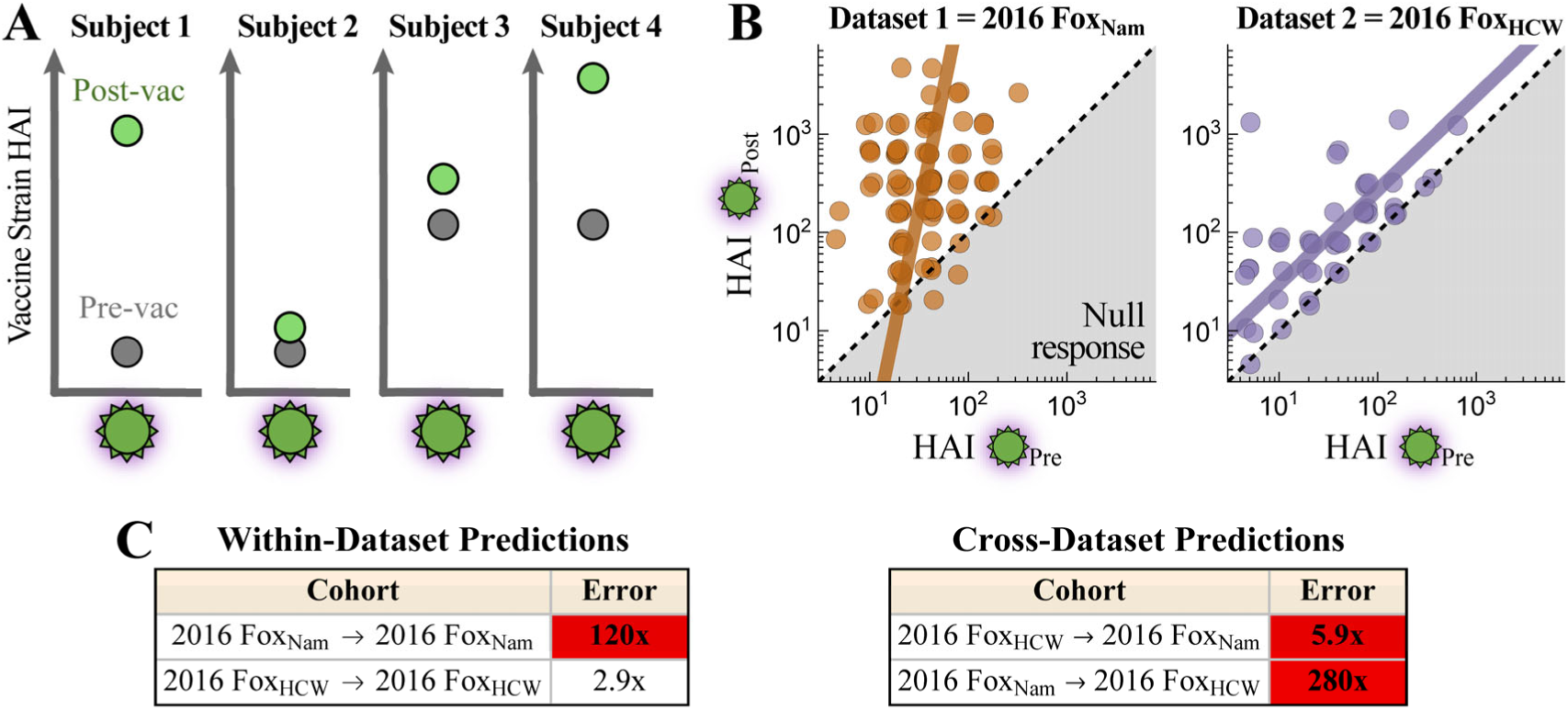
Using pre-vaccination titers alone can lead to large prediction error across studies. Comparison of vaccine strain (H3N2 A/Hong Kong/4801/2014) HAIs across the 2016 Fox_Nam_ and 2016 Fox_HCW_ studies. (A) Schematic emphasizing that two people may have the same HAI pre-vac (HAI_pre_) but end up with very different HAI post-vac (HAI_post_). (B) Pre– and post-vac HAI for the vaccine strain across both studies. There is heterogeneity both within one study (distribution of points around each best-fit line) as well as heterogeneity across studies (differences between the best-fit lines). (C) For within-dataset predictions of the vaccine strain’s pre→post-vac HAI, linear regression was fit to 70% of the data to predict the remaining 30%, with error showing the mean fold-change across 10 iterations. For cross-dataset predictions, all data from one cohort was used to predict all data in the other cohort. Cases where error>4x are highlighted in red. Note that some errors reach >100x.

**Figure S9.**
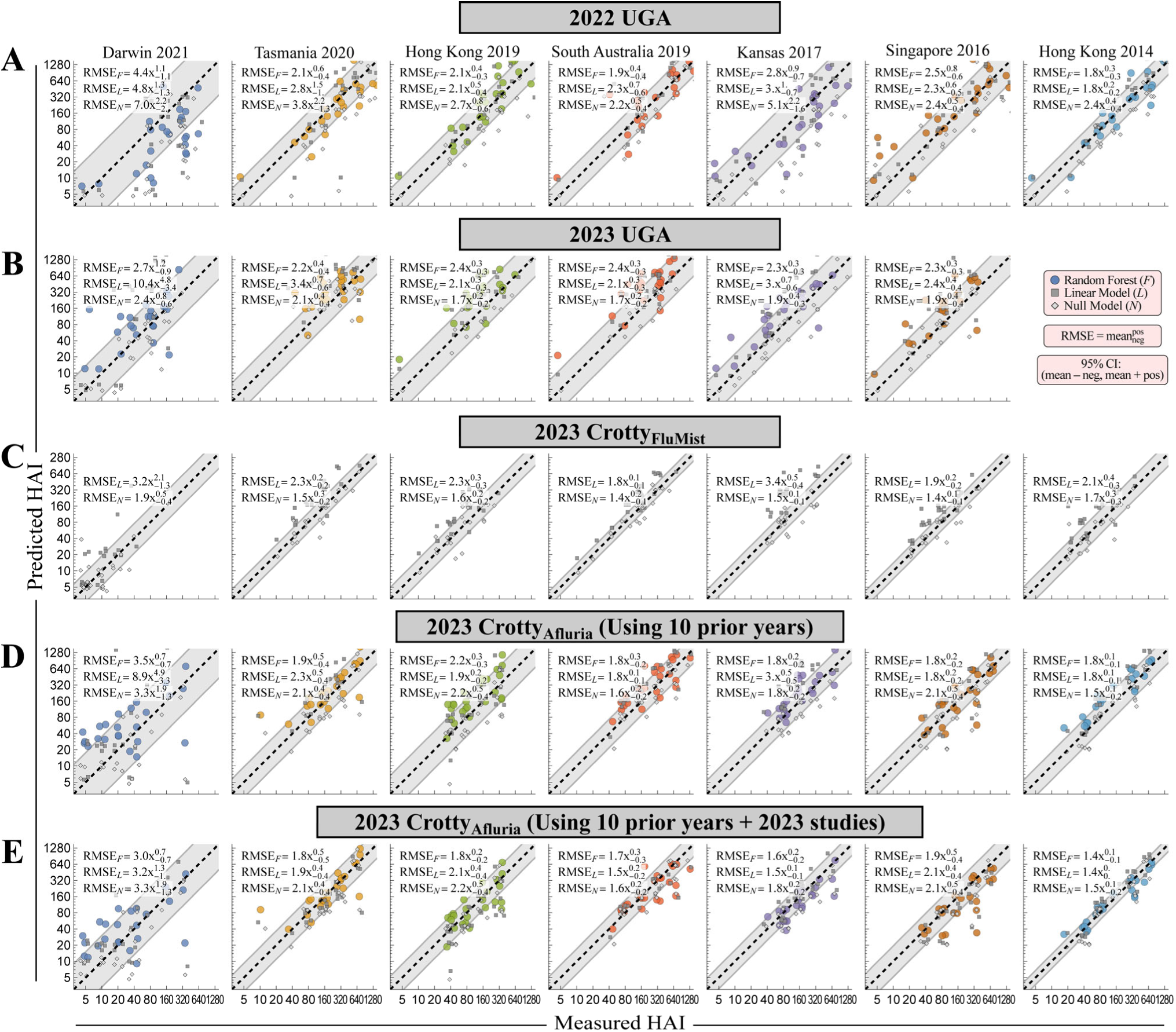
Individual virus predictions from. Fig 4. Predicted versus measured post-vac HAI for the new vaccine studies (A) 2022 UGA, (B) 2023 UGA [which did not measure Hong Kong 2014], (C) 2023 Crotty_FluMist_, and (D) 2023 Crotty_Afluria_ using the random forest approach (*F*, colored points), linear model (*L*, gray squares), or null model (*N*, gray diamonds). For all predictions, training was carried out on studies from the past 10 years; random forests were not used for the live attenuated vaccine study (2023 Crotty_FluMist_). RMSE and 95% confidence intervals (sub/superscripts) are shown in the top-left. (E) Since the 2023 Crotty_Afluria_ study was conducted three months after the 2023 UGA study, an additional specialized prediction was created by finding the best combination of vaccine studies that predicted the 2023 UGA data and then using those to predict 2023 Crotty_Afluria_. The resulting predictions were based upon 2022 and 2023 vaccine studies [some currently unpublished]. The gray diagonal band denotes the width of RMSE_F_ in panels A,B,D,E and RMSE_N_ in panel C.

**Figure S10.**
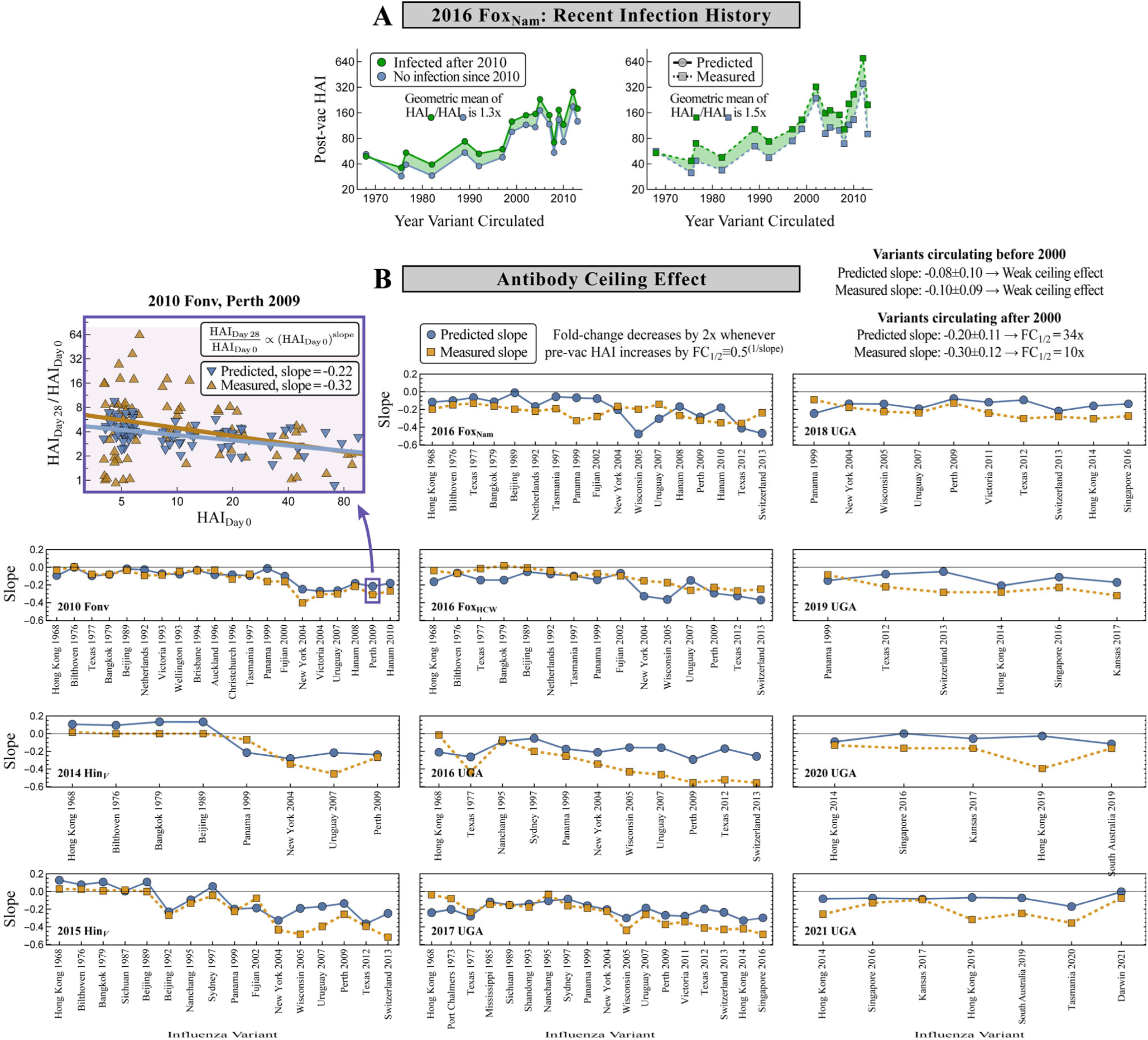
Recouping the effects of recent infection history and the antibody ceiling. (A) Predicted versus measured post-vac HAI for the 2016 Fox_Nam_ study, showing the geometric mean of the responses for n=51 individuals with a known influenza H3N2 infection after 2010 (green) and n=49 individuals with last confirmed H3N2 infection before 2010 (blue). The shaded area indicates that the more recently infected cohort has a larger HAI. (B) The antibody ceiling effect was computed for each variant and every study using the slope of the linear fit of pre-vac HAI versus fold-change (both on log-axes). Resulting slopes are shown for all measurements and predictions.

**Figure S11.**
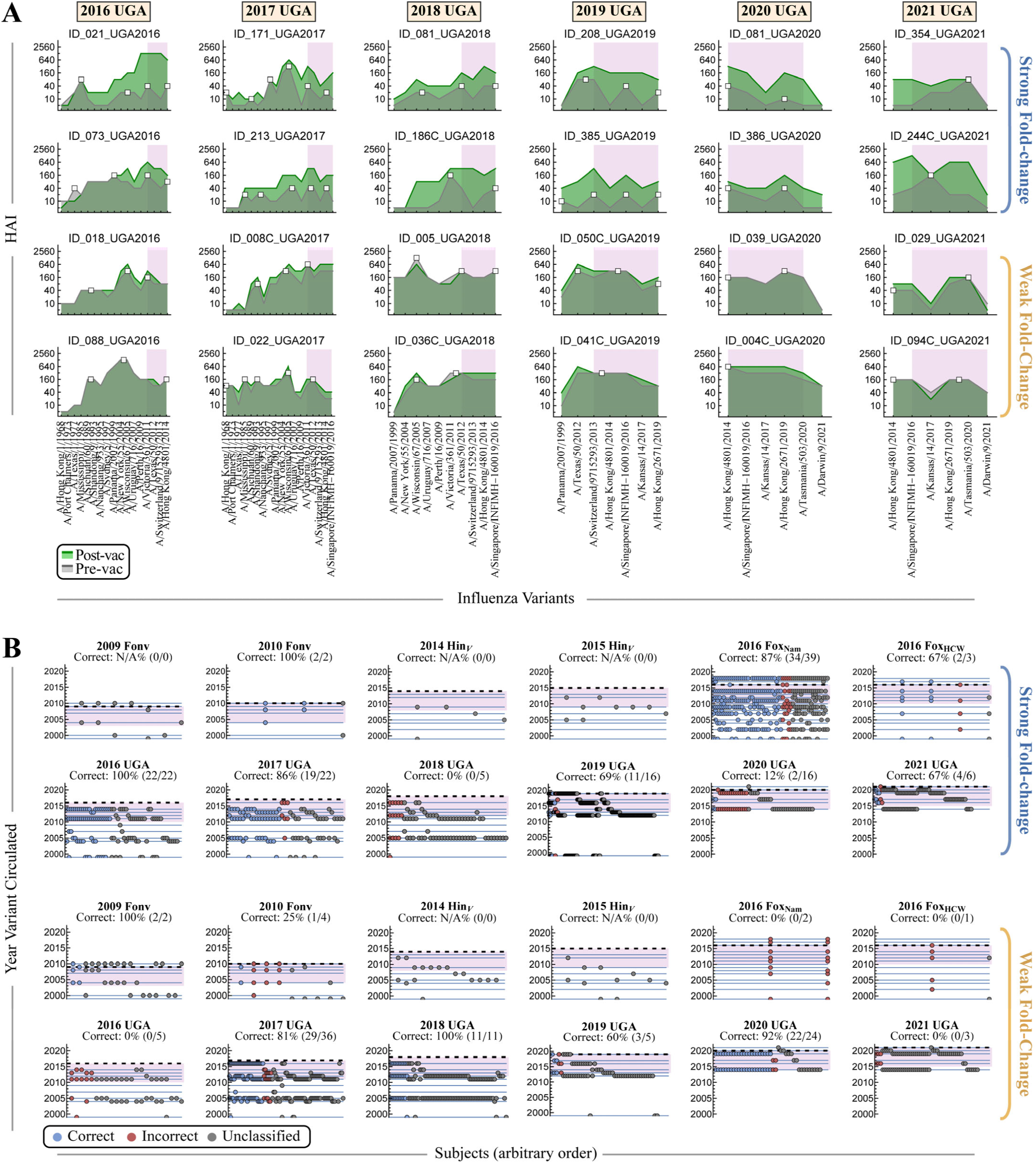
Characterizing “robustly strong” and “robustly weak” vaccine responses. (A) Representative examples of robust strong or weak responses from the UGA studies. A support vector machine and nearest-neighbors classified each state, as well as ≥85% of perturbed states (varying any single HAI by 2x or 1/2x), as this same strong/weak state. (B) Classifying subjects with strong or weak fold-change post-vaccination from all studies. Each subject is either correctly classified (blue), incorrectly classified (red), or not classified (gray). Blue lines represent the years of all variants from 2000 onwards in each study, and points on each column represent the peaks of an individual’s HAI profile. In Panels A and B, the purple region represents the region in which Δ_Peak_ is computed (*i.e.*, the period 0-6 years before each study). The dashed line in Panel B represents the year of each study.

**Figure S12.**
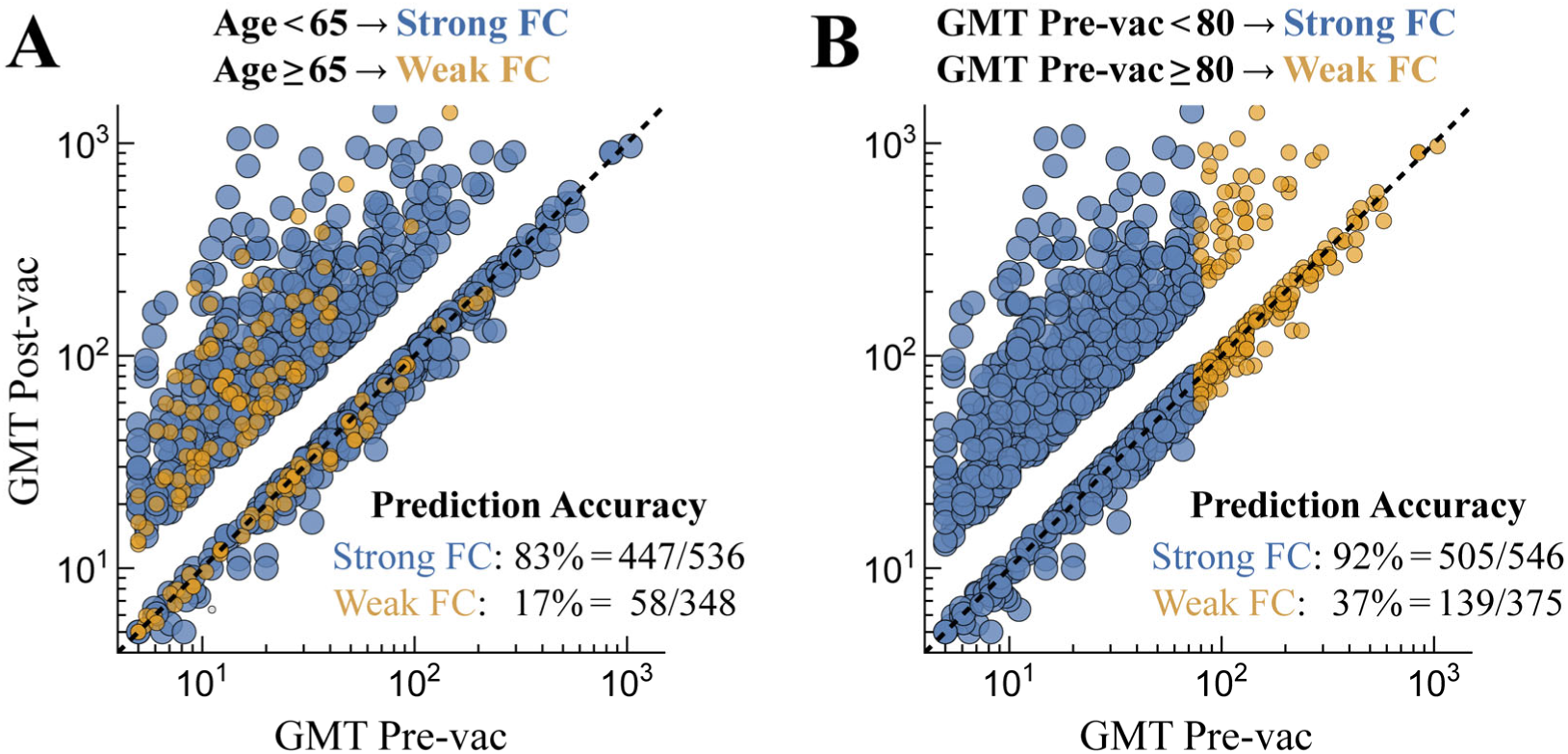
Alternate categorization of strong/weak fold-change responses. (A) An age-based categorization assuming the elderly (age≥65) exhibit weak fold-change (FC) post-vaccination. (B) Pre-vac GMT-based categorization assuming that individuals that start with higher titers (GMT pre-vac≥80) will exhibit weak fold-change. In both panels, statistics in the bottom-right show how many of the actual strong responses (FC≥2.5) or weak responses (FC≤1.1) were correctly predicted.

## References

1. Sun, H., Yang, J., Zhang, T., Long, L.-P., Jia, K., Yang, G., Webby, R.J., and Wan, X.-F. (2013). Using Sequence Data To Infer the Antigenicity of Influenza Virus. mBio 4, e00230–00213-e00230. 10.1128/mbio.00230-13.

2. Petrova, V.N., and Russell, C.A. (2018). The evolution of seasonal influenza viruses. Nat Rev Microbiol 16, 47–60. 10.1038/nrmicro.2017.118.

3. Morris, D.H., Gostic, K.M., Pompei, S., Bedford, T., Luksza, M., Neher, R.A., Grenfell, B.T., Lassig, M., and McCauley, J.W. (2018). Predictive Modeling of Influenza Shows the Promise of Applied Evolutionary Biology. Trends Microbiol 26, 102–118. 10.1016/j.tim.2017.09.004.

4. Fonville, J.M., Wilks, S.H., James, S.L., Fox, A., Ventresca, M., Aban, M., Xue, L., Jones, T.C., Le, N.M.H., Pham, Q.T., et al. (2014). Antibody landscapes after influenza virus infection or vaccination. Science 346, 996–1000. 10.1126/science.1256427.

5. Cobey, S., and Hensley, S.E. (2017). Immune history and influenza virus susceptibility. Curr Opin Virol 22, 105–111. 10.1016/j.coviro.2016.12.004.

6. McLean, H.Q., Thompson, M.G., Sundaram, M.E., Meece, J.K., McClure, D.L., Friedrich, T.C., and Belongia, E.A. (2014). Impact of repeated vaccination on vaccine effectiveness against influenza A(H3N2) and B during 8 seasons. Clin Infect Dis 59, 1375–1385. 10.1093/cid/ciu680.

7. Lewnard, J.A., and Cobey, S. (2018). Immune History and Influenza Vaccine Effectiveness. Vaccines (Basel) 6. 10.3390/vaccines6020028.

8. Jang, H., and Ross, T.M. (2019). Preexisting influenza specific immunity and vaccine effectiveness. Expert Review of Vaccines 18, 1043–1051. 10.1080/14760584.2019.1675519.

9. Auladell, M., Phuong, H.V.M., Mai, L.T.Q., Tseng, Y.Y., Carolan, L., Wilks, S., Thai, P.Q., Price, D., Duong, N.T., Hang, N.L.K., et al. (2022). Influenza virus infection history shapes antibody responses to influenza vaccination. Nat Med 28, 363–372. 10.1038/s41591-022-01690-w.

10. Zhao, X., Fang, V.J., Ohmit, S.E., Monto, A.S., Cook, A.R., and Cowling, B.J. (2016). Quantifying Protection Against Influenza Virus Infection Measured by Hemagglutination-inhibition Assays in Vaccine Trials. Epidemiology 27, 143–151. 10.1097/EDE.0000000000000402.

11. Cowling, B.J., Lim, W.W., Perera, R., Fang, V.J., Leung, G.M., Peiris, J.S.M., and Tchetgen Tchetgen, E.J. (2019). Influenza Hemagglutination-inhibition Antibody Titer as a Mediator of Vaccine-induced Protection for Influenza B. Clin Infect Dis 68, 1713–1717. 10.1093/cid/ciy759.

12. Krammer, F. (2019). The human antibody response to influenza A virus infection and vaccination. Nat Rev Immunol 19, 383–397. 10.1038/s41577-019-0143-6.

13. Coudeville, L., Bailleux, F., Riche, B., Megas, F., Andre, P., and Ecochard, R. (2010). Relationship between haemagglutination-inhibiting antibody titres and clinical protection against influenza: development and application of a bayesian random-effects model. BMC Medical Research Methodology 10, 18. 10.1186/1471-2288-10-18.

14. Allison, M.A., Daley, M.F., Crane, L.A., Barrow, J., Beaty, B.L., Allred, N., Berman, S., and Kempe, A. (2006). Influenza vaccine effectiveness in healthy 6-to 21-month-old children during the 2003-2004 season. The Journal of Pediatrics 149, 755–762.e751. 10.1016/j.jpeds.2006.06.036.

15. Rudenko, L., Naykhin, A., Donina, S., Korenkov, D., Petukhova, G., Isakova-Sivak, I., Losev, I., Stukova, M., Erofeeva, M., Nikiforova, A., et al. (2015). Assessment of immune responses to H5N1 inactivated influenza vaccine among individuals previously primed with H5N2 live attenuated influenza vaccine. Human Vaccines & Immunotherapeutics 11, 2839–2848. 10.1080/21645515.2015.1069931.

16. Lu, X., Liu, F., Tzeng, W.-P., York, I.A., Tumpey, T.M., and Levine, M.Z. (2024). Antibody-Mediated Suppression Regulates the Humoral Immune Response to Influenza Vaccination in Humans. The Journal of Infectious Diseases 229, 310–321. 10.1093/infdis/jiad493.

17. Goodwin, E., Gibbs, J.S., Yewdell, J.W., Eisenlohr, L.C., and Hensley, S.E. (2024). Influenza virus antibodies inhibit antigen-specific de novo B cell responses in mice. bioRxiv. 10.1101/2024.04.12.589218.

18. Lessler, J., Riley, S., Read, J.M., Wang, S., Zhu, H., Smith, G.J., Guan, Y., Jiang, C.Q., and Cummings, D.A. (2012). Evidence for antigenic seniority in influenza A (H3N2) antibody responses in southern China. PLoS Pathog 8, e1002802. 10.1371/journal.ppat.1002802.

19. Yewdell, J.W., and Santos, J.J.S. (2021). Original Antigenic Sin: How Original? How Sinful? Cold Spring Harb Perspect Med 11. 10.1101/cshperspect.a038786.

20. Oidtman, R.J., Arevalo, P., Bi, Q., McGough, L., Russo, C.J., Vera Cruz, D., Costa Vieira, M., and Gostic, K.M. (2021). Influenza immune escape under heterogeneous host immune histories. Trends Microbiol 29, 1072–1082. 10.1016/j.tim.2021.05.009.

21. Brouwer, A.F., Balmaseda, A., Gresh, L., Patel, M., Ojeda, S., Schiller, A.J., Lopez, R., Webby, R.J., Nelson, M.I., Kuan, G., and Gordon, A. (2022). Birth cohort relative to an influenza A virus’s antigenic cluster introduction drives patterns of children’s antibody titers. PLoS Pathog 18, e1010317. 10.1371/journal.ppat.1010317.

22. Gostic, K.M., Ambrose, M., Worobey, M., and Lloyd-Smith, J.O. (2016). Potent protection against H5N1 and H7N9 influenza via childhood hemagglutinin imprinting. Science 354, 722–726. 10.1126/science.aag1322.

23. Vinh, D.N., Nhat, N.T.D., De Bruin, E., Vy, N.H.T., Thao, T.T.N., Phuong, H.T., Anh, P.H., Todd, S., Quan, T.M., Thanh, N.T.L., et al. (2021). Age-seroprevalence curves for the multi-strain structure of influenza A virus. Nature Communications 12. 10.1038/s41467-021-26948-8.

24. Avey, S., Mohanty, S., Chawla, D.G., Meng, H., Bandaranayake, T., Ueda, I., Zapata, H.J., Park, K., Blevins, T.P., Tsang, S., et al. (2020). Seasonal Variability and Shared Molecular Signatures of Inactivated Influenza Vaccination in Young and Older Adults. The Journal of Immunology 204, 1661–1673. 10.4049/jimmunol.1900922.

25. Diray-Arce, J., Miller, H.E.R., Henrich, E., Gerritsen, B., Mulè, M.P., Fourati, S., Gygi, J., Hagan, T., Tomalin, L., Rychkov, D., et al. (2022). The Immune Signatures data resource, a compendium of systems vaccinology datasets. Scientific Data 9. 10.1038/s41597-022-01714-7.

26. Ranjeva, S., Subramanian, R., Fang, V.J., Leung, G.M., Ip, D.K.M., Perera, R., Peiris, J.S.M., Cowling, B.J., and Cobey, S. (2019). Age-specific differences in the dynamics of protective immunity to influenza. Nat Commun 10, 1660. 10.1038/s41467-019-09652-6.

27. Forst, C.V., Chung, M., Hockman, M., Lashua, L., Adney, E., Hickey, A., Carlock, M., Ross, T., Ghedin, E., and Gresham, D. (2022). Vaccination History, Body Mass Index, Age, and Baseline Gene Expression Predict Influenza Vaccination Outcomes. Viruses 14. 10.3390/v14112446.

28. Tsang, J.S., Schwartzberg, P.L., Kotliarov, Y., Biancotto, A., Xie, Z., Germain, R.N., Wang, E., Olnes, M.J., Narayanan, M., Golding, H., et al. (2014). Global analyses of human immune variation reveal baseline predictors of postvaccination responses. Cell 157, 499–513. 10.1016/j.cell.2014.03.031.

29. Cortese, M., Sherman, A.C., Rouphael, N.G., and Pulendran, B. (2021). Systems Biological Analysis of Immune Response to Influenza Vaccination. Cold Spring Harbor Perspectives in Medicine 11, a038596. 10.1101/cshperspect.a038596.

30. Einav, T., and Cleary, B. (2022). Extrapolating missing antibody-virus measurements across serological studies. Cell Syst 13, 561–573 e565. 10.1016/j.cels.2022.06.001.

31. Einav, T., and Ma, R. (2023). Using interpretable machine learning to extend heterogeneous antibody-virus datasets. Cell Rep Methods 3, 100540. 10.1016/j.crmeth.2023.100540.

32. Hodgson, D., Sanchez-Ovando, S., Carolan, L., Liu, Y., Hadiprodjo, A.J., Fox, A., Sullivan, S.G., and Kucharski, A.J. (2024). Quantifying the impact of pre-vaccination titre and vaccination history on influenza vaccine immunogenicity. medRxiv. 10.1101/2024.01.24.24301614.

33. Hobson, D., Curry, R.L., Beare, A.S., and Ward-Gardner, A. (1972). The role of serum haemagglutination-inhibiting antibody in protection against challenge infection with influenza A2 and B viruses. Epidemiology and Infection 70, 767–777. 10.1017/s0022172400022610.

34. Paccalin, M., Plouzeau, C., Bouche, G., Guillard, O., Beby-Defaux, A., Mauco, G., and Agius, G. (2006). Lack of correlation between nutritional status and seroprotection against influenza in a long term care facility. Scandinavian Journal of Infectious Diseases 38, 894–897. 10.1080/00365540600749984.

35. Black, S., Nicolay, U., Vesikari, T., Knuf, M., Del Giudice, G., Della Cioppa, G., Tsai, T., Clemens, R., and Rappuoli, R. (2011). Hemagglutination Inhibition Antibody Titers as a Correlate of Protection for Inactivated Influenza Vaccines in Children. Pediatric Infectious Disease Journal 30, 1081–1085. 10.1097/inf.0b013e3182367662.

36. Hay, J.A., Zhu, H., Jiang, C.Q., Kwok, K.O., Shen, R., Kucharski, A., Yang, B., Read, J.M., Lessler, J., Cummings, D.A.T., and Riley, S. (2024). Reconstructed influenza A/H3N2 infection histories reveal variation in incidence and antibody dynamics over the life course. bioRxiv. 10.1101/2024.03.18.24304371.

37. Hinojosa, M., Shepard, S.S., Chung, J.R., King, J.P., McLean, H.Q., Flannery, B., Belongia, E.A., and Levine, M.Z. (2021). Impact of Immune Priming, Vaccination, and Infection on Influenza A(H3N2) Antibody Landscapes in Children. J Infect Dis 224, 469–480. 10.1093/infdis/jiaa665.

38. Fox, A., Carolan, L., Leung, V., Phuong, H.V.M., Khvorov, A., Auladell, M., Tseng, Y.Y., Thai, P.Q., Barr, I., Subbarao, K., et al. (2022). Opposing Effects of Prior Infection versus Prior Vaccination on Vaccine Immunogenicity against Influenza A(H3N2) Viruses. Viruses 14. 10.3390/v14030470.

39. Carlock, M.A., Allen, J.D., Hanley, H.B., and Ross, T.M. (2024). Longitudinal assessment of human antibody binding to hemagglutinin elicited by split-inactivated influenza vaccination over six consecutive seasons. PLOS ONE 19, e0301157. 10.1371/journal.pone.0301157.

40. Henry, C., Zheng, N.Y., Huang, M., Cabanov, A., Rojas, K.T., Kaur, K., Andrews, S.F., Palm, A.E., Chen, Y.Q., Li, Y., et al. (2019). Influenza Virus Vaccination Elicits Poorly Adapted B Cell Responses in Elderly Individuals. Cell Host Microbe 25, 357–366 e356. 10.1016/j.chom.2019.01.002.

41. Gouma, S., Kim, K., Weirick, M.E., Gumina, M.E., Branche, A., Topham, D.J., Martin, E.T., Monto, A.S., Cobey, S., and Hensley, S.E. (2020). Middle-aged individuals may be in a perpetual state of H3N2 influenza virus susceptibility. Nat Commun 11, 4566. 10.1038/s41467-020-18465-x.

42. Welsh, F.C., Eguia, R.T., Lee, J.M., Haddox, H.K., Galloway, J., Van Vinh Chau, N., Loes, A.N., Huddleston, J., Yu, T.C., Quynh Le, M., et al. (2024). Age-dependent heterogeneity in the antigenic effects of mutations to influenza hemagglutinin. Cell Host & Microbe. 10.1016/j.chom.2024.06.015.

43. Nakaya, H.I., Hagan, T., Duraisingham, S.S., Lee, E.K., Kwissa, M., Rouphael, N., Frasca, D., Gersten, M., Mehta, A.K., and Gaujoux, R. (2015). Systems analysis of immunity to influenza vaccination across multiple years and in diverse populations reveals shared molecular signatures. Immunity 43, 1186–1198. 10.1016%2Fj.immuni.2015.11.012

44. Avey, S., Cheung, F., Fermin, D., Frelinger, J., Gaujoux, R., Gottardo, R., Khatri, P., Kleinstein, S.H., Kotliarov, Y., Meng, H., et al. (2017). Multicohort analysis reveals baseline transcriptional predictors of influenza vaccination responses. Science Immunology 2, eaal4656. 10.1126/sciimmunol.aal4656.

45. Andrews, S.F., Huang, Y., Kaur, K., Popova, L.I., Ho, I.Y., Pauli, N.T., Dunand, C.J.H., Taylor, W.M., Lim, S., Huang, M., et al. (2015). Immune history profoundly affects broadly protective B cell responses to influenza. Science Translational Medicine 7, 316ra192–316ra311. 10.1126/scitranslmed.aad0522.

46. Hopping, A.M., McElhaney, J., Fonville, J.M., Powers, D.C., Beyer, W.E.P., and Smith, D.J. (2016). The confounded effects of age and exposure history in response to influenza vaccination. Vaccine 34, 540–546. 10.1016/j.vaccine.2015.11.058.

47. Loes, A.N., Tarabi, R.A.L., Huddleston, J., Touyon, L., Wong, S.S., Cheng, S.M.S., Leung, N.H.L., Hannon, W.W., Bedford, T., Cobey, S., et al. (2024). High-throughput sequencing-based neutralization assay reveals how repeated vaccinations impact titers to recent human H1N1 influenza strains. bioRxiv. 10.1101/2024.03.08.584176.

48. Koel, B.F., Burke, D.F., Bestebroer, T.M., Van Der Vliet, S., Zondag, G.C.M., Vervaet, G., Skepner, E., Lewis, N.S., Spronken, M.I.J., Russell, C.A., et al. (2013). Substitutions Near the Receptor Binding Site Determine Major Antigenic Change During Influenza Virus Evolution. Science 342, 976–979. 10.1126/science.1244730.

49. Barría, M.I., Garrido, J.L., Stein, C., Scher, E., Ge, Y., Engel, S.M., Kraus, T.A., Banach, D., and Moran, T.M. (2013). Localized Mucosal Response to Intranasal Live Attenuated Influenza Vaccine in Adults. The Journal of Infectious Diseases 207, 115–124. 10.1093/infdis/jis641.

50. Williams, K.V., Zhai, B., Alcorn, J.F., Patricia Nowalk, M., Levine, M.Z., Kim, S.S., Flannery, B., Moehling Geffel, K., Jaber Merranko, A., Nagg, J.P., et al. (2022). A randomized controlled trial of antibody response to 2019–20 cell-based inactivated and egg-based live attenuated influenza vaccines in children and young adults. Vaccine 40, 780–788. 10.1016/j.vaccine.2021.12.034.

51. Yegorov, S., Celeste, D.B., Gomes, K.B., Ang, J.C., Vandenhof, C., Wang, J., Rybkina, K., Tsui, V., Stacey, H.D., Loeb, M., and Miller, M.S. (2022). Inactivated and live-attenuated seasonal influenza vaccines boost broadly neutralizing antibodies in children. Cell Reports Medicine 3, 100509. 10.1016/j.xcrm.2022.100509.

52. Meade, P., Strohmeier, S., Bermúdez-González, M.C., García-Sastre, A., Palese, P., Simon, V., and Krammer, F. (2023). Antigenic Landscape Analysis of Individuals Vaccinated with a Universal Influenza Virus Vaccine Candidate Reveals Induction of Cross-Subtype Immunity. Journal of Virology 97. 10.1128/jvi.01070-22.

53. Hoft, D.F., Babusis, E., Worku, S., Spencer, C.T., Lottenbach, K., Truscott, S.M., Abate, G., Sakala, I.G., Edwards, K.M., Creech, C.B., et al. (2011). Live and Inactivated Influenza Vaccines Induce Similar Humoral Responses, but Only Live Vaccines Induce Diverse T-Cell Responses in Young Children. The Journal of Infectious Diseases 204, 845–853. 10.1093/infdis/jir436.

54. Block, S.L., Falloon, J., Hirschfield, J.A., Krilov, L.R., Dubovsky, F., Yi, T., and Belshe, R.B. (2012). Immunogenicity and Safety of a Quadrivalent Live Attenuated Influenza Vaccine in Children. Pediatric Infectious Disease Journal 31, 745–751. 10.1097/inf.0b013e31825687b0.

55. Manenti, A., Tete, S.M., Mohn, K.G.I., Jul-Larsen, Å., Gianchecchi, E., Montomoli, E., Brokstad, K.A., and Cox, R.J. (2017). Comparative analysis of influenza A(H3N2) virus hemagglutinin specific IgG subclass and IgA responses in children and adults after influenza vaccination. Vaccine 35, 191–198. 10.1016/j.vaccine.2016.10.024.

56. Ertesvag, N.U., Cox, R.J., Lartey, S.L., Mohn, K.G., Brokstad, K.A., and Trieu, M.C. (2022). Seasonal influenza vaccination expands hemagglutinin-specific antibody breadth to older and future A/H3N2 viruses. NPJ Vaccines 7, 67. 10.1038/s41541-022-00490-0.

57. Block, S.L., Yi, T., Sheldon, E., Dubovsky, F., and Falloon, J. (2011). A randomized, double-blind noninferiority study of quadrivalent live attenuated influenza vaccine in adults. Vaccine 29, 9391–9397. 10.1016/j.vaccine.2011.09.109.

58. Yang, B., Garcia-Carreras, B., Lessler, J., Read, J.M., Zhu, H., Metcalf, C.J.E., Hay, J.A., Kwok, K.O., Shen, R., Jiang, C.Q., et al. (2022). Long term intrinsic cycling in human life course antibody responses to influenza A(H3N2): an observational and modeling study. Elife 11. 10.7554/eLife.81457.

59. Meijers, M., Ruchnewitz, D., Eberhardt, J., Karmakar, M., Łuksza, M., and Lässig, M. (2024). Concepts and methods for predicting viral evolution. bioRxiv. 10.1101/2024.03.19.585703.

60. Kucharski, A.J., Lessler, J., Read, J.M., Zhu, H., Jiang, C.Q., Guan, Y., Cummings, D.A.T., and Riley, S. (2015). Estimating the Life Course of Influenza A(H3N2) Antibody Responses from Cross-Sectional Data. PLOS Biology 13, e1002082. 10.1371/journal.pbio.1002082.

61. Kucharski, A.J., Lessler, J., Cummings, D.A.T., and Riley, S. (2018). Timescales of influenza A/H3N2 antibody dynamics. PLoS Biol 16, e2004974. 10.1371/journal.pbio.2004974.

62. Kim, K., Vieira, M.C., Gouma, S., Weirick, M.E., Hensley, S.E., and Cobey, S. (2024). Measures of population immunity can predict the dominant clade of influenza A (H3N2) in the 2017-2018 season and reveal age-associated differences in susceptibility and antibody-binding specificity. medRxiv. 10.1101/2023.10.26.23297569.

63. Nachbagauer, R., Choi, A., Hirsh, A., Margine, I., Iida, S., Barrera, A., Ferres, M., Albrecht, R.A., Garcia-Sastre, A., Bouvier, N.M., et al. (2017). Defining the antibody cross-reactome directed against the influenza virus surface glycoproteins. Nat Immunol 18, 464–473. 10.1038/ni.3684.

64. Allen, J.D., and Ross, T.M. (2021). Evaluation of Next-Generation H3 Influenza Vaccines in Ferrets Pre-Immune to Historical H3N2 Viruses. Front Immunol 12, 707339. 10.3389/fimmu.2021.707339.

65. Uno, N., and Ross, T.M. (2024). Multivalent next generation influenza virus vaccines protect against seasonal and pre-pandemic viruses. Scientific Reports 14. 10.1038/s41598-023-51024-0.

66. Ge, Y., Lu, Y., Allen, J., Einav, T., Nkaleke, D.I., Bai, F., Handel, A., Ross, T., and Shen, Y. (2024). Pre-existing Immunity to Influenza Aids Ferrets in Developing Stronger and Broader Vaccine-induced Antibody Responses. Vaccine.

67. DiazGranados, C.A., Dunning, A.J., Kimmel, M., Kirby, D., Treanor, J., Collins, A., Pollak, R., Christoff, J., Earl, J., Landolfi, V., et al. (2014). Efficacy of High-Dose versus Standard-Dose Influenza Vaccine in Older Adults. New England Journal of Medicine 371, 635–645. 10.1056/nejmoa1315727.

68. Sanchez, L., Nakama, T., Nagai, H., Matsuoka, O., Inoue, S., Inoue, T., Shrestha, A., Pandey, A., Chang, L.-J., and De Bruijn, I. (2023). Superior immunogenicity of high-dose quadrivalent inactivated influenza vaccine versus Standard-Dose vaccine in Japanese Adults ≥60 years of age: Results from a phase III, randomized clinical trial. Vaccine 41, 2553–2561. 10.1016/j.vaccine.2023.02.071.

69. Johansen, N.D., Modin, D., Skaarup, K.G., Nealon, J., Samson, S., Dufournet, M., Loiacono, M.M., Harris, R.C., Larsen, C.S., Jensen, A.M.R., et al. (2024). Effectiveness of high-dose versus standard-dose quadrivalent influenza vaccine against recurrent hospitalizations and mortality in relation to influenza circulation: A post-hoc analysis of the DANFLU-1 randomized clinical trial. Clinical Microbiology and Infection. 10.1016/j.cmi.2024.01.017.

70. Vardeny, O., Kim, K., Udell, J.A., Joseph, J., Desai, A.S., Farkouh, M.E., Hegde, S.M., Hernandez, A.F., McGeer, A., Talbot, H.K., et al. (2021). Effect of High-Dose Trivalent vs Standard-Dose Quadrivalent Influenza Vaccine on Mortality or Cardiopulmonary Hospitalization in Patients With High-risk Cardiovascular Disease. JAMA 325, 39. 10.1001/jama.2020.23649.

71. Wu, S., Ross, T.M., Carlock, M.A., Ghedin, E., Choi, H., and Vogel, C. (2022). Evaluation of determinants of the serological response to the quadrivalent split-inactivated influenza vaccine. Molecular Systems Biology 18. 10.15252/msb.202110724.

72. CDC (2024). Influenza Fact Sheet. https://www.cdc.gov/flu/about/keyfacts.htm.

73. Harvey, W.T., Benton, D.J., Gregory, V., Hall, J.P., Daniels, R.S., Bedford, T., Haydon, D.T., Hay, A.J., McCauley, J.W., and Reeve, R. (2016). Identification of Low– and High-Impact Hemagglutinin Amino Acid Substitutions That Drive Antigenic Drift of Influenza A(H1N1) Viruses. PLoS Pathog 12, e1005526. 10.1371/journal.ppat.1005526.

